# Prothrombotic antiphospholipid antibodies in COVID-19

**DOI:** 10.1101/2020.06.15.20131607

**Authors:** Yu Zuo, Shanea K. Estes, Ramadan A. Ali, Alex A. Gandhi, Srilakshmi Yalavarthi, Hui Shi, Gautam Sule, Kelsey Gockman, Jacqueline A. Madison, Melanie Zuo, Vinita Yadav, Jintao Wang, Wrenn Woodward, Sean P. Lezak, Njira L. Lugogo, Stephanie A. Smith, James H. Morrissey, Yogendra Kanthi, Jason S. Knight

## Abstract

Patients with coronavirus disease 19 (**COVID-19**) are at high risk for thrombotic arterial and venous occlusions. At the same time, lung histopathology often reveals fibrin-based occlusion in the small vessels of patients who succumb to the disease. Antiphospholipid syndrome (**APS**) is an acquired and potentially life-threatening thrombophilia in which patients develop pathogenic autoantibodies (**aPL**) targeting phospholipids and phospholipid-binding proteins. Case series have recently detected aPL in patients with COVID-19. Here, we measured eight types of aPL [anticardiolipin IgG/IgM/IgA, anti-beta-2 glycoprotein I IgG/IgM/IgA, and anti-phosphatidylserine/prothrombin (**aPS/PT**) IgG/IgM] in the sera of 172 patients hospitalized with COVID-19. We detected aPS/PT IgG in 24%, anticardiolipin IgM in 23%, and aPS/PT IgM in 18%. Any aPL was present in 52% of patients using the manufacturer’s threshold and in 30% using a more stringent cutoff (≥40 units). Higher levels of aPL were associated with neutrophil hyperactivity (including the release of neutrophil extracellular traps/**NETs**), higher platelet count, more severe respiratory disease, and lower glomerular filtration rate. Similar to patients with longstanding APS, IgG fractions isolated from patients with COVID-19 promoted NET release from control neutrophils. Furthermore, injection of these COVID-19 IgG fractions into mice accelerated venous thrombosis. Taken together, these studies suggest that a significant percentage of patients with COVID-19 become at least transiently positive for aPL and that these aPL are potentially pathogenic.

## INTRODUCTION

Abnormal coagulation parameters correlate with COVID-19 severity (*1, 2*). D-dimer, in particular, is an independent risk factor for death (*1, 3-5*). Early descriptions of the COVID-19 coagulopathy identified it as disseminated intravascular coagulation. However, most patients maintain normal levels of coagulation factors, fibrinogen, and platelets suggesting that COVID-19 induces a unique prothrombotic state that is distinct from traditional descriptions of sepsis-induced coagulopathy (*6, 7*). There are now increasing reports of venous thromboembolism in patients with COVID-19 (*8, 9*). This observation is despite concerns regarding under-diagnosis given baseline elevations in the sensitive biomarker D-dimer, as well as pragmatic challenges in obtaining diagnostic imaging while patients are in isolation. Arterial thrombosis including strokes and myocardial infarctions have also been described (*9, 10*). Histopathology of lung specimens from patients with severe disease show not only characteristic findings of ARDS but also evidence of fibrin-based occlusion of small vessels (*11-13*). There are several (possibly synergistic) mechanisms by which SARS-CoV-2 infection may result in macrovascular and microvascular thrombosis (*14*), including a cytokine storm that activates leukocytes, endothelium, and platelets; hypoxic vaso-occlusion; and direct activation of cells by viral transduction. Furthermore, many patients hospitalized with COVID-19 have high levels of neutrophil extracellular traps (**NETs**) in their blood (*15, 16*) and these inflammatory cell remnants may also contribute to the prothrombotic milieu (*17-20*).

Antiphospholipid syndrome (**APS**) is an acquired thrombophilia, affecting at least 1 in 2000 individuals (*21*). Patients form durable autoantibodies to phospholipids and phospholipid-binding proteins such as prothrombin and beta-2-glycoprotein I (**β_2_GPI**). These autoantibodies engage cell surfaces, where they activate endothelial cells, platelets, and neutrophils (*22, 23*), thereby tipping the blood-endothelium interface toward thrombosis. A key feature of APS is its ability to promote thrombosis in vascular beds of all sizes, including both arterial and venous circuits. The catastrophic variant of APS is frequently fatal, and bears some similarities to the diffuse coagulopathy seen in patients with COVID-19 (*24*). Classification criteria for APS (last updated in 2006) seek persistently positive testing for anticardiolipin (**aCL**) or anti-β_2_GPI antibodies (**aβ_2_GPI**) (*25*). Lupus anticoagulant testing (a functional assay that screens for aPL based on their paradoxical ability to prolong *in vitro* clotting assays) is also included in the criteria and detects a variety of species of aPL including anti-phosphatidylserine/prothrombin (**aPS/PT**) (*26*).

Reports of antiphospholipid antibodies (**aPL**) in COVID-19 and their possible relationship to thrombosis have begun to emerge in case reports and series (*27-32*). While viral infections are well-known triggers of transient aPL (*33-36*), the extent to which these short-lived antibodies are pathogenic has never been well defined. Here, we endeavored to test several types of aPL in a large cohort of patients hospitalized with COVID-19. We also asked whether purified IgG fractions from these patients had prothrombotic properties *in vitro* and *in vivo*.

## RESULTS

### Prevalence of aPL in patients hospitalized with COVID-19

Sera from 172 patients hospitalized with COVID-19 (**Supplementary Table 1**) were evaluated for eight different types of aPL. Of the 172 patients, 19% died and 8% remained in the hospital at the time of this analysis. Eighty-nine patients tested positive for at least one type of aPL based on the manufacturer’s cut-off, representing 52% of the entire cohort (**Table 1**). Lupus anticoagulant testing was not performed given lack of access to fresh plasma samples. Among the various aPL tested, aPS/PT IgG had the highest prevalence (24%), followed by aCL IgM (23%) and aPS/PT IgM (18%). Forty-one patients (24%) were positive for more than one type of aPL and 13 (8%) were positive for more than two. Fifty-two patients (30%) had at least one moderate- to high-titer aPL (**Table 1**). Thirty-six patients had sera from multiple time points available for aPL testing and those longitudinal results are presented in **Supplementary Figure 1**; the highest aPL levels were used to classify positivity for these 36 patients in the aforementioned analysis (**Table 1**). Seropositivity was also assessed using only the first available sample with similar results (**Supplementary Table 2**). To further elucidate the antigen specificity of patients with positive aPS/PT antibodies, we also measured anti-phosphatidylserine (**aPS**) antibodies in sera that were positive for aPS/PT. Neither aPS IgG nor aPS IgM correlated with aPS/PT levels suggesting prothrombin-dependence of aPS/PT antibodies (**Supplementary Figure 2**). In summary, any positive aPL was detected in 52% of patients hospitalized for COVID-19, with approximately two-thirds of those being detected at moderate-to-high titers. The majority of positives were associated with three antibodies: aPS/PT IgG, aCL IgM, and aPS/PT IgM.

**Table 1:**
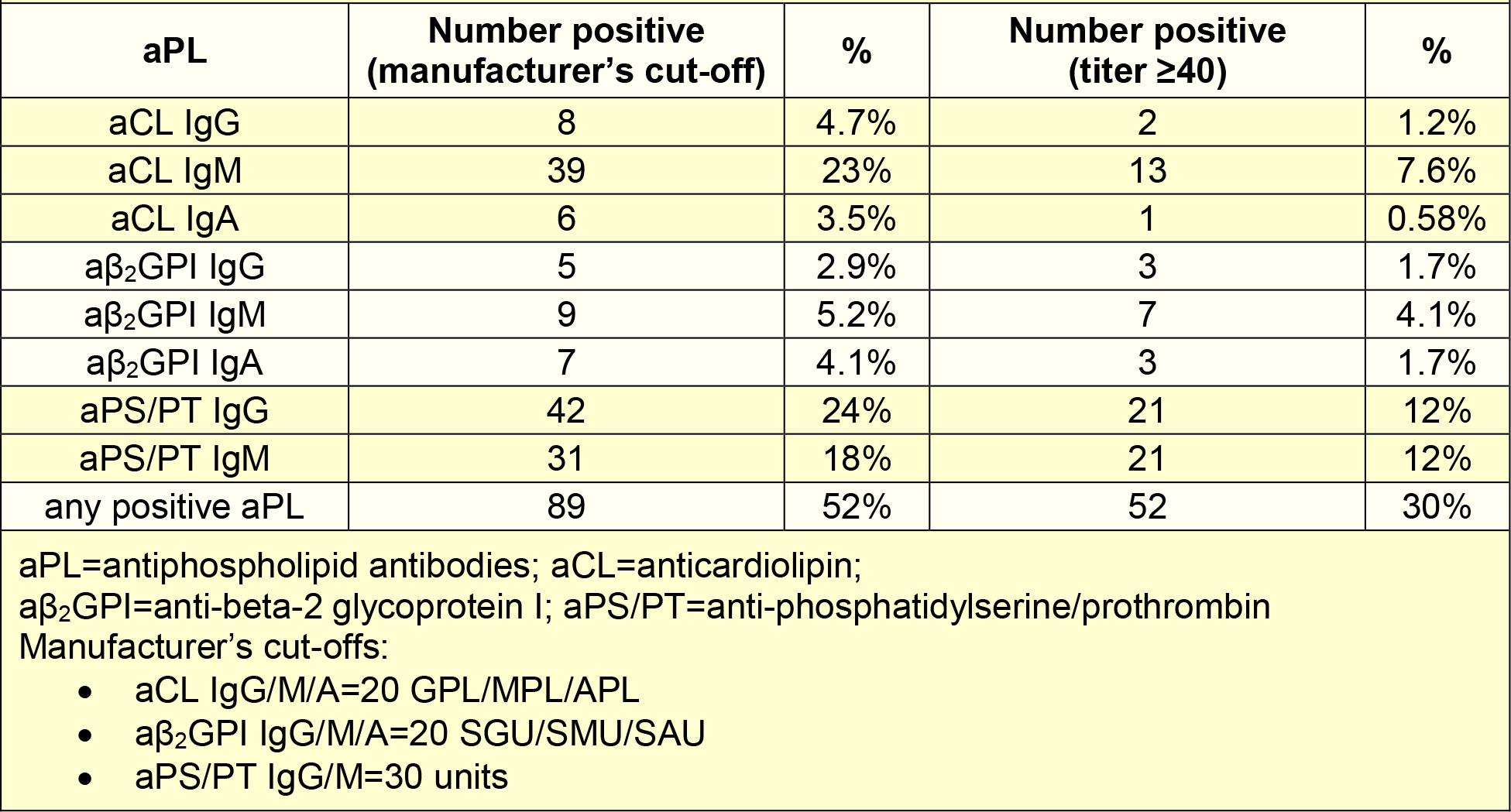
Prevalence of antiphospholipid antibodies in COVID-19 patients (n=172)

### Clinical correlations of COVID-19-associated aPL

We next asked whether the presence of aPL associated with various clinical parameters. Specifically, we assessed potential correlations with SpO_2_/FiO_2_ ratio (i.e., oxygenation efficiency), C-reactive protein, D-dimer, platelet count, absolute neutrophil count, calprotectin (a marker of neutrophil activation), and myeloperoxidase-DNA complexes (a marker of NETs) (**Table 2**). Levels of anticardiolipin IgM were significantly correlated with all of the aforementioned parameters (**Table 2**). Neutrophil activation (calprotectin) most consistently associated with various different aPL (**Table 2**). We also assessed a previously devised scoring tool (**aPL-S**), which integrates and prioritizes data from the various aPL types tested (see Methods) (*37*). aPL-S demonstrated a positive correlation with platelet count, neutrophil activation, and NETs (**Table 2**). We then examined various clinical parameters as they related to positive/negative aPL thresholds. Positive testing for any aPL was associated with higher levels of calprotectin and lower estimated glomerular filtration rate (**eGFR**) (**Figure 1A-B**). These differences were also observed when comparing patients with positive aPS/PT to the remainder of the cohort (**Figure 1C-D**) or to patients without any aPL (**Supplementary Figure 3**). Nadir eGFR was also lower in patients with a history of renal disease as compared to those without (**Supplementary Figure 4**). Oxygenation efficiency tended to be impaired in patients with positive aPL or aPS/PT testing as compared to those without, although group comparisons did not reach statistical significance (**Supplementary Figure 5**). Similarly, peak troponin (for any aPL) and peak D-dimer (for aPS/PT) tended to be higher in patients with positive testing (**Supplementary Figure 6**). Since obesity can affect the D-dimer levels, we compared D-dimer in patients with and without obesity, but did not find a difference (**Supplementary Figure 7**). Taken together, COVID-19-associated aPL track with various clinical parameters, especially neutrophil activation and impaired renal function.

**Fig. 1:**
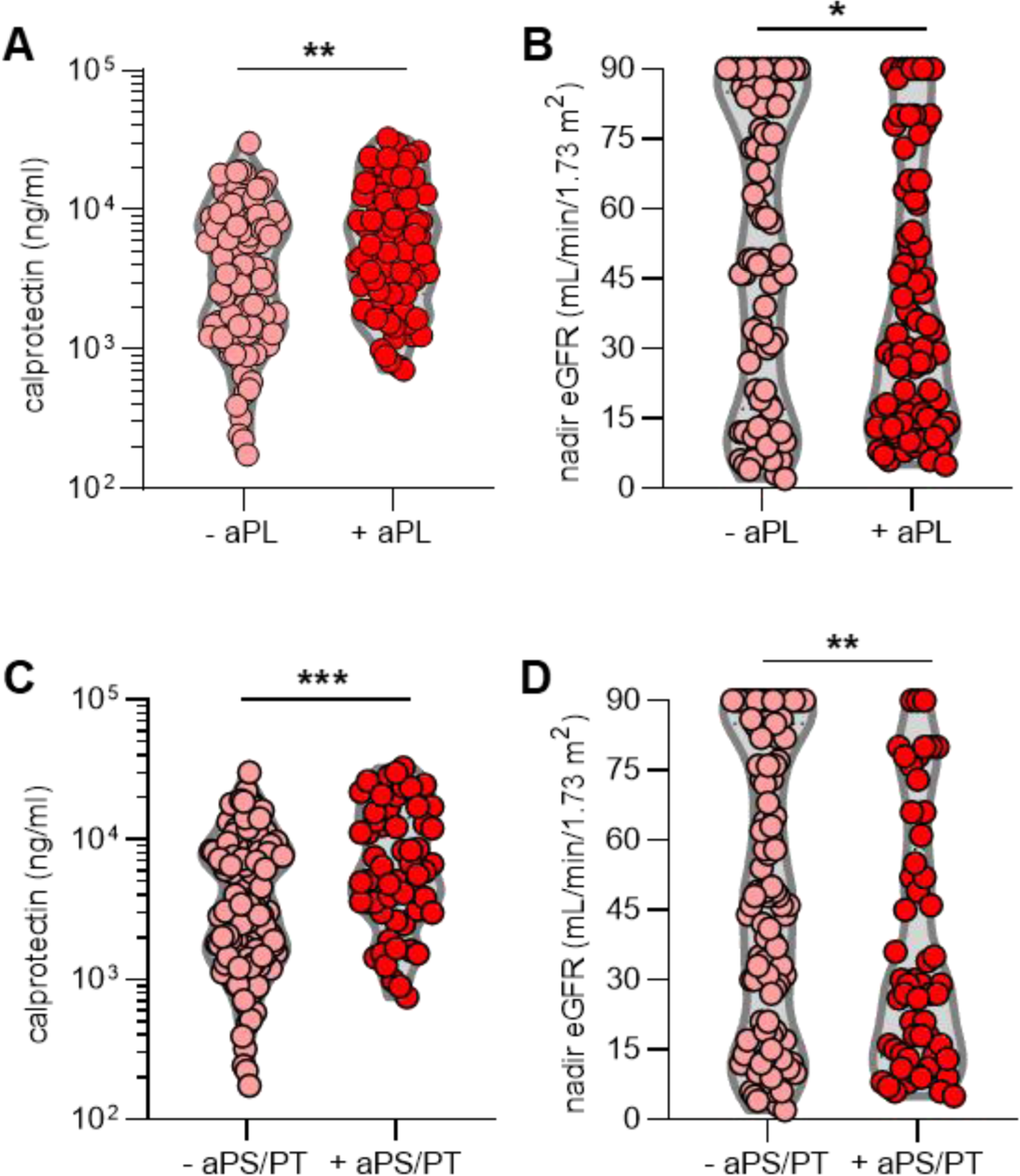
Positive testing for aPL is associated with more neutrophil activation and worse kidney function. **A-B**, N=172 COVID-19 patients were divided into groups based on positivity (manufacturer’s threshold) for any aPL. **C-D**, COVID-19 patients were grouped based on positive testing for aPS/PT (IgG and IgM considered together); manufacturer’s thresholds were used to determine positivity. Groups were analyzed by unpaired t-test; *p<0.05, **p<0.01, and ***p<0.001. eGFR=estimated glomerular filtration rate. For patients who had data available from multiple time points, only the first-available sample was used in this analysis.

**Table 2:** Correlation of antiphospholipid antibodies with clinical and laboratory variables in COVID-19 patients**‡**.

|  |  |  |  |  |  |  |  |  |  |  |  |  |  |  |
| --- | --- | --- | --- | --- | --- | --- | --- | --- | --- | --- | --- | --- | --- | --- |
|  | aPL-S score<br>(modified) |  | aCL IgG |  | aCL IgM |  | aβ<sub>2</sub>GPI IgG |  | aβ<sub>2</sub>GPI IgM |  | aPS/PT IgG |  | aPS/PT IgM |  |
| Spearman | r | p | r | p | r | p | r | p | r | p | r | p | r | p |
| Clinical and lab variables |  |  |  |  |  |  |  |  |  |  |  |  |  |  |
| SpO2/FiO2 | -0.051 | ns | -0.16 | * | -0.19 | * | -0.10 | ns | -0.022 | ns | -0.11 | ns | -0.16 | * |
| C-reactive protein | 0.031 | ns | 0.15 | ns | 0.17 | * | 0.075 | ns | -0.040 | ns | 0.058 | ns | 0.16 | * |
| D-dimer | 0.087 | ns | 0.092 | ns | 0.24 | ** | 0.041 | ns | 0.000 | ns | 0.005 | ns | 0.037 | ns |
| Platelet count | 0.17 | * | 0.095 | ns | 0.29 | **** | 0.17 | * | 0.11 | ns | -0.009 | ns | 0.23 | ** |
| Neutrophil count | 0.10 | ns | 0.13 | ns | 0.19 | * | 0.047 | ns | 0.041 | ns | -0.008 | ns | 0.096 | ns |
| Calprotectin | 0.26 | *** | 0.29 | **** | 0.28 | *** | 0.11 | ns | 0.090 | ns | 0.25 | *** | 0.23 | ** |
| NETs (MPO/DNA) | 0.18 | * | 0.16 | * | 0.25 | *** | 0.20 | ** | 0.13 | ns | 0.033 | ns | 0.23 | ** |
| ns=not significant; NETs=neutrophil extracellular traps; MPO=myeloperoxidase; *p<0.05, **p<0.01, ***p<0.001, and **** p<0.0001 |  |  |  |  |  |  |  |  |  |  |  |  |  |  |
| ‡ 36 patients had sera from multiple time points; for those patients only the first available sample was used for determining correlations. |  |  |  |  |  |  |  |  |  |  |  |  |  |  |

### IgG isolated from COVID-19 patients triggers NET release

Work by our group and others has revealed that one prothrombotic function of aPL in patients with APS is to trigger NET release (*22, 38*). Given that we recently detected elevated levels of NETs in COVID-19 patients (*15*), we reasoned that IgG fractions purified from patients with COVID-19 might be able to trigger NET release. We selected two COVID-19 patients with high aβ_2_GPI IgG, two with high aPS/PT IgG, and two without any positive aPL. From these patients, we purified total IgG fractions and tested them alongside IgG pooled from two patients with active catastrophic antiphospholipid syndrome (**CAPS**), as well as a separate pool from five patients with longstanding triple-positive APS (positive testing for aCL, aβ_2_GPI, and lupus anticoagulant). The purity of isolated COVID-19 IgG was verified by SDS-PAGE (**Supplementary Figure 8**). As compared with unstimulated neutrophils, COVID-19 IgG samples positive for aPL doubled NET release, similar to CAPS and triple-positive APS samples (**Figure 2A**). Representative COVID-19 IgG-induced NETs are shown in **Figure 2B**. Work by our group has previously shown that dipyridamole—an antithrombotic medication—can attenuate aPL-mediated prothrombotic NET release via surface adenosine A_2A_ receptor agonism (*39*). Here, we found that dipyridamole also suppresses COVID-19 IgG-mediated NET release (**Supplementary Figure 9**). In summary, IgG fractions purified from aPL-positive COVID-19 patients promote NET release similar to IgG isolated from individuals with established APS.

**Fig. 2:**
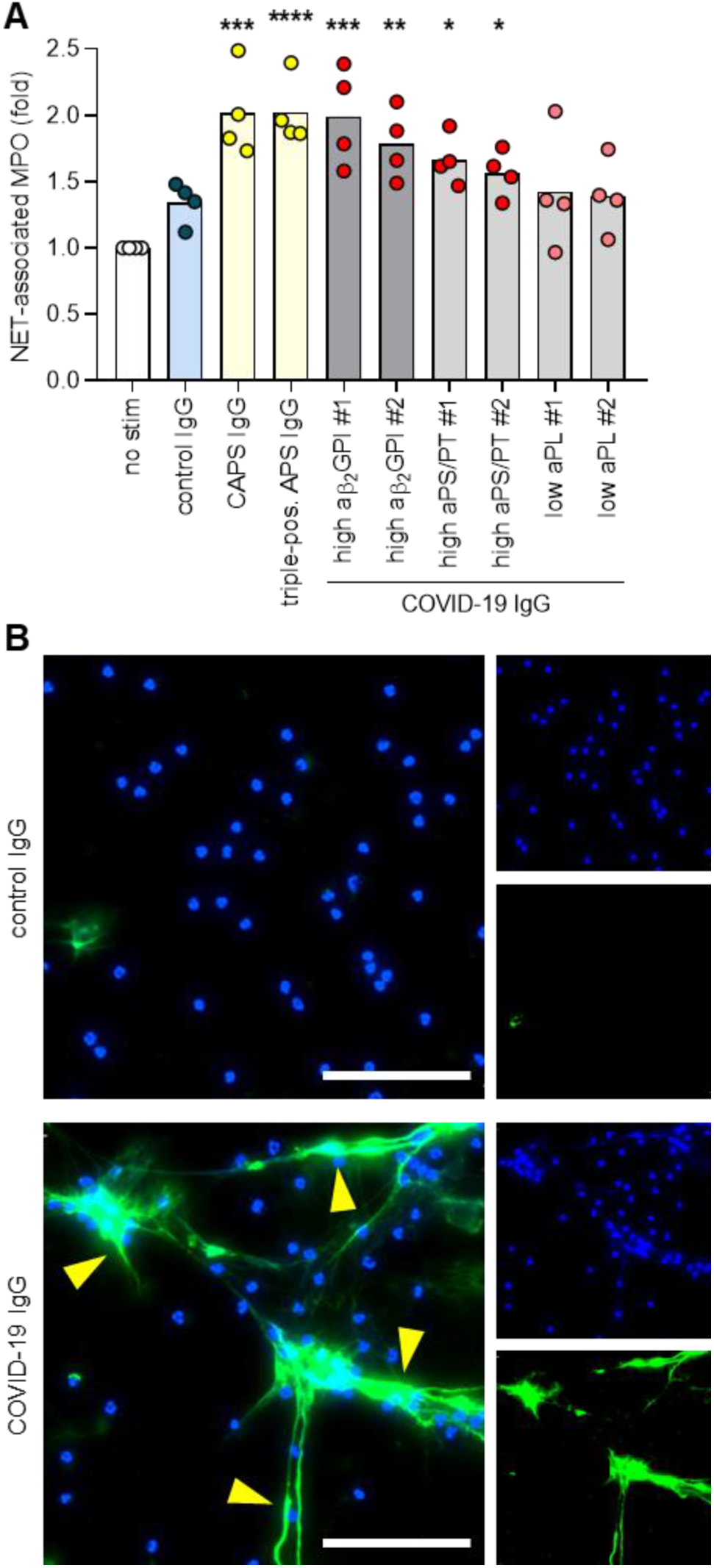
APS and COVID IgG promote the release of neutrophil extracellular traps from control neutrophils. **A**, Neutrophils were isolated from healthy controls and cultured in the presence of human IgG (10 μg/ml) as indicated for 3 hours. NET release was measured by the enzymatic activity of myeloperoxidase (MPO) after solubilization of NETs with Micrococcal nuclease. Data are presented for four independent experiments. Comparisons were by one-way ANOVA with correction for multiple comparisons by Dunnett’s method. **B**, Representative microscopy with DNA stained blue and neutrophil elastase green. Some examples of NETs are indicated with yellow arrowheads. Scale bars=100 microns.

### aPL-positive COVID-19 IgG potentiates thrombosis *in vivo*

We next sought to determine whether IgG fractions from COVID-19 patients could accelerate thrombosis. When tested in an *in vitro* cell-free thrombin generation assay, IgG fractions purified from COVID-19 patients did not have demonstrable clot-accelerating activity (**Supplementary Figure 10**); nevertheless, we speculated that a prothrombotic phenotype might still be appreciated in the cell-enriched vascular environment *in vivo*. We have previously reported that IgG isolated from patients with either CAPS or triple-positive APS accelerates large-vein thrombosis in various mouse models of inferior vena cava thrombosis (*38-40*). Here, we asked whether COVID-19 IgG might behave similarly to enhance thrombosis under such conditions. In a model that activates the endothelium by local electrolysis-mediated free radicals (**Figure 3A**), IgG isolated from patients with high levels of aPS/PT IgG increased thrombus extension (Figure 3B) and overall accretion (**Figure 3C-D**). At the same time, the high aPS/PT samples significantly increased circulating NET remnants, similar to CAPS IgG (**Figure 3E**), and demonstrated a tendency toward high levels of thrombus citrullinated histone H3 (a biochemical marker of NETs) (**Supplementary Figure 11**). To confirm these findings, we turned our attention to an independent model in which the IVC is narrowed just distal to the renal veins by a fixed suture (**Figure 3F**). In this “stenosis” model, a high aPS/PT COVID-19 IgG preparation again increased thrombus extension (**Figure 3G**), thrombus accretion (**Figure 3H-I**), and circulating NET remnants (**Figure 3J**). Taken together, these data indicate that IgG fractions from some patients with acute COVID-19 are able to accelerate thrombosis *in vivo*.

**Fig. 3:**
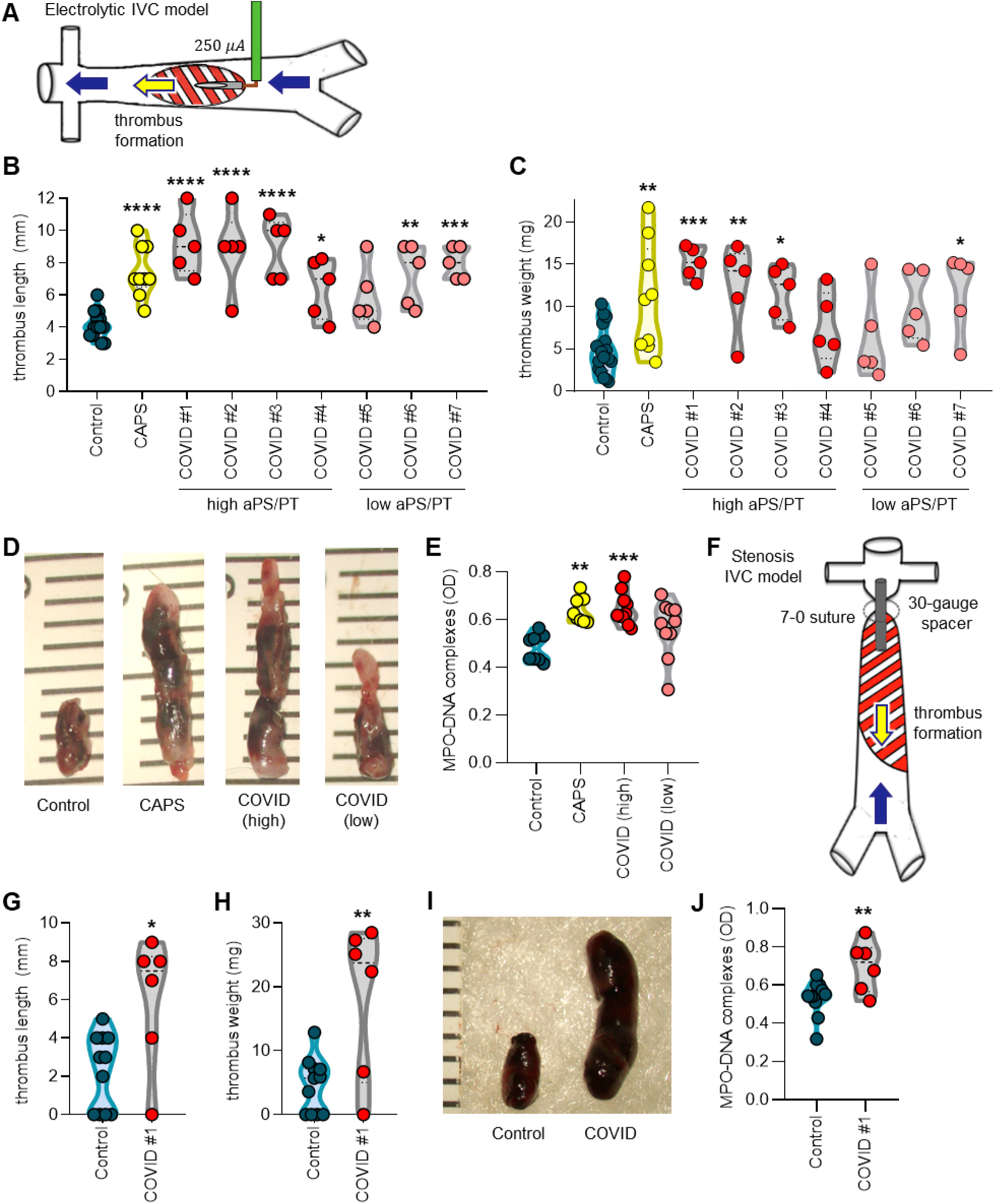
COVID-19 IgG potentiates thrombosis *in vivo*. **A**, Model of thrombus initiation in the inferior vena cava (IVC) by local electrolysis. **B-C**, Mice were treated with control, CAPS, or COVID-19 IgG as indicated and thrombus size was determined 24 hours after local electrolysis; each point represents an individual mouse. **D**, Representative thrombi from B-C. **E**, Sera from some mice included in B-C were tested for NET remnants as measured by myeloperoxidase-DNA ELISA. **F**, Model of thrombus initiation in the IVC by fixed stenosis. **G-H**, Mice were treated with COVID-19 IgG as indicated and thrombus size was determined 24 hours after stenosis. **I**, Representative thrombi from G-H. **J**, Sera from some mice included in G-H were tested for NET remnants. Comparisons were by either one-way ANOVA with correction for multiple comparisons by Dunnett’s method (B, C, E) or unpaired t test (G, H, J); *p<0.05, **p<0.01, ***p<0.001, and ****p<0.0001.

## DISCUSSION

aPL are a heterogeneous group of autoantibodies that underlie the pathogenesis of APS via their interactions with phospholipid-binding plasma proteins such as β_2_GPI, prothrombin, thrombomodulin, plasminogen, antithrombin III, protein C, protein S, annexin II, annexin V, and likely others (*41-47*). The association between various infections and the induction of aPL has long been recognized (*48-53*). For example, one study of 100 cases reported in Medline from 1983 to 2003 found the most commonly reported aPL-associated infections to be skin infections (18%), pneumonia (14%), and urinary tract infections (10%); common organisms included human immunodeficiency virus (17%), varicella-zoster virus (15%), and hepatitis C virus (13%) (51). Regarding specific antibodies, aCL IgG and IgM (typically lacking anti-β_2_GPI activity) have been most commonly reported (*49, 53-58*). The majority of these virus-associated aPL are thought to be transient (35, *55, 59)*. While the clinical significance of transient virus-associated aPL remains to be fully defined, a recent review of 163 published cases of virus-associated aPL found thrombotic events in 116 (*35*). While acknowledging the likelihood of sampling and publication bias, these data (along with the data presented here for individuals with relatively severe COVID-19) suggest that some transient aPL may still have prothrombotic potential. Whether similar findings would extend to patients with less symptomatic COVID-19 presentations—some of whom may experience thrombotic events—awaits further study.

The most severe presentation of APS is its catastrophic variant known as CAPS, which fortunately impacts only a minority of APS patients—typically at times of stress such as infection, surgery, or anticoagulation withdrawal (*60*). CAPS involves derangements of both inflammatory and thrombotic pathways and impacts multiple organs in the body simultaneously (*60*). In the largest series of CAPS patients assembled, the most commonly affected organs were kidneys (73%), lungs (60%), brain (56%), heart (50%), and skin (47%) (*61*). While multiorgan failure may certainly complicate severe cases of COVID-19, the lungs are typically the most severely affected. We speculate that local immune stimulation from viral infection (including potentially the infection of endothelial cells) could synergize with circulating aPL and thereby lead to a particularly severe thromboinflammatory insult to the lungs of COVID-19 patients.

Many studies from the general thrombosis literature have revealed that activated neutrophils, and in particular NET formation, contribute to the propagation of thrombi affecting arterial, venous, and microscopic vascular beds (*62, 63*). NETs have also been recently implicated in the pathogenesis of APS. In 2015, our group reported that sera from APS patients, as well as purified aPL, trigger neutrophils to release NETs (*22*). The potential *in vivo* relevance of this observation has been confirmed in mouse models of aPL-mediated large-vein thrombosis in which either depletion of neutrophils or digestion of NETs is protective (*38*). Neutrophils from APS patients also appear to have increased adhesive potential, which is dependent upon the activated form of integrin Mac-1; this pro-adhesive phenotype amplifies neutrophil-endothelium interactions, potentiates NET formation, and potentially lowers the threshold for thrombosis (*64*). Therapies that target NET formation have the potential to treat thrombotic diseases. For example, selective agonism of the adenosine A_2A_ receptor suppresses aPL-mediated NETosis in protein kinase A-dependent fashion (*39*). A_2A_ agonism also reduces thrombosis in the inferior vena cava of both control mice and mice treated with aPL. Dipyridamole, which is known to potentiate adenosine receptor signaling by increasing extracellular concentrations of adenosine and interfering with the breakdown of cAMP, also suppresses aPL-mediated NETosis and mitigates venous thrombosis in mice (*65*). Interestingly, a small study from China showed that dipyridamole significantly suppressed D-dimer elevation and improved platelet counts in patients with COVID-19 (*66*). While we have demonstrated here that dipyridamole mitigated COVID-19 IgG-mediated NET release, prospective randomized clinical trials (NCT04391179) are needed to evaluate clinical outcomes among COVID-19 patients (*67*).

“Criteria” aPL are defined based on their inclusion in the updated Sapporo classification criteria: namely, aCL IgG and IgM, aβ_2_GPI IgG and IgM, and lupus anticoagulant (*25*); of these, lupus anticoagulant is generally accepted as the best indicator of a high-risk aPL profile (*68-73*). At the same time, there are certainly reports of patients with “seronegative APS,” who have classic features of APS but with negative testing for criteria aPL (*74*). Some non-criteria aPL discovered in the past 20 years have shown promising clinical utility in identifying APS. Among those are aPS/PT IgG and IgM, as well as the IgA isotypes of aCL and aβ_2_GPI. Retrospective studies have suggested that aβ_2_GPI IgA antibodies are significantly associated with thrombosis in lupus patients (OR 2.8, 95% CI 1.3-6.2) (75). A recent review of 10 retrospective studies (1775 patients with lupus or primary APS and 628 healthy controls) detected a strong association between aPS/PT and thrombotic events (OR 5.11; 95% CI 4.2-6.3) (*76*). Furthermore, serological agreement between aPS/PT IgG/IgM and high-risk aPL profiles—especially the presence of lupus anticoagulant—has been demonstrated in a recent study of 95 well-characterized primary APS patients (*77*). While the clinical significance of aPS/PT during viral infection remains to be comprehensively defined, we found here that IgG fractions containing high levels of these antibodies trigger NET release *in vitro* and accelerate thrombosis *in vivo*. It is also interesting to note that COVID-19 IgG purified from low-aPS/PT patients demonstrated some activity in potentiating thrombosis (although high-aPS/PT fractions were more robust). It is possible that aPL are but one species of a broader acute natural antibody response that is in fact prothrombotic in COVID-19.

The orchestration of autoimmunity against phospholipids in COVID-19 is likely a complex interplay between genetic predisposition, historical antigen exposures, and a hyperactivated host immune response in the setting of a unique environmental trigger—SARS-CoV-2 (*78*). It is probably not surprising that aPL of the IgM isotype (which are designed for rapid mobilization) predominate in this cohort where they correlate with markers of neutrophil activation and NET release. The relationship between aPL and NETs in COVID-19 is potentially bidirectional. NETs are a known source of autoantigens, while cytokines released in parallel to NETosis may also facilitate NET-associated autoantibody propagation (*79-82*). An example of a cytokine that could play such a role is B-cell activating factor (BAFF), an important mediator of B-cell maturation into antibody-producing cells (*83*). For example, neutrophil-derived BAFF likely participates in the production of anti-double-stranded DNA antibodies in lupus (*80*). In COVID-19, it is possible that production of aPL potentiates NET formation and BAFF release. This may further enhance the survival and differentiation of phospholipid-reactive B cells, and in some cases class-switching to the IgG isotype. The interplay between COVID-19 and humoral immunity is clearly an area that merits further study.

There are several potential clinical implications of these findings. Patients with CAPS are regularly treated with heparin, corticosteroids, and plasmapheresis—with the addition of the latter leading to a demonstrable improvement in outcomes (*84*). While both anticoagulation and corticosteroids have shown some promise to date in treatment of COVID-19, plasmapheresis has not been systematically explored. One wonders if this could provide benefit in the subgroup of COVID-19 patients with high titers of aPL. At the same time, convalescent plasma is receiving increasing attention as an approach to treating severe cases of COVID-19. Defining the extent to which these samples may contain aPL or other autoantibodies, in addition to protective anti-SARS-CoV-2 antibodies, is another potential area for future investigation.

This study has some limitations. We did not have access to the fresh plasma that would be required for lupus anticoagulant testing (which would have provided additional context and risk stratification to the aPL profiling). We speculate that many of the patients with positive aPS/PT would have displayed a lupus anticoagulant phenotype—as reported recently (*26*)—but proving that will require further study and prospective access to samples. We were also not able to define a clear link between circulating aPL and large artery/vein thrombosis. Eleven patients in our cohort had thrombotic events and 55% of those were positive for aPL. It should be noted that aggressive anticoagulation has been regularly employed at our institution including many patients treated prophylactically with therapeutic doses of anticoagulation. It should also be noted that aPL were not tested on a defined hospital day, but rather when a sample became available to the research laboratory; future studies should endeavor to systematically track aPL over the full course of a hospitalization, and perhaps especially at and after the time of discharge. In the meantime, and as we await definitive antiviral and immunologic solutions to the current pandemic, we posit that testing aPL, including aPS/PT, may lead to improved risk stratification and personalization of treatment for COVID-19, and that aPL are at least a part of the complex COVID-19 thromboinflammatory milieu.

## MATERIALS AND METHODS

### Human samples

Serum samples from 172 hospitalized COVID-19 patients were used in this study (**Supplementary Table 1**). Blood was collected into serum separator tubes containing clot activator and serum separator gel by a trained hospital phlebotomist. After completion of biochemical testing ordered by the clinician, the remaining serum was stored for clinical testing at 4°C for up to 48 hours before release to the research laboratory. Serum samples were immediately divided into small aliquots and stored at −80°C until the time of testing. All 172 patients had a confirmed COVID-19 diagnosis based on FDA-approved RNA testing. Fifty of these 172 patients were included in our prior study evaluating the role of NETs in COVID-19 (*85*), although aPL were not considered in that study (*85*). This study complied with all relevant ethical regulations, and was approved by the University of Michigan Institutional Review Board (HUM00179409), which waived the requirement for informed consent given the discarded nature of the samples.

### Quantification of aPL

aPL were quantified in sera using Quanta Lite® ACA IgG, ACA IgM, ACA IgA, β_2_GPI IgG, β_2_GPI IgM, β_2_GPI IgA, aPS IgG, aPS IgM, aPS/PT IgG, and aPS/PT IgM kits (Inova Diagnostics Inc.) according to the manufacturer’s instructions. All assays are approved for clinical use and received 510(k) clearance from the FDA. Quanta Lite® aPL ELISAs (Inova Diagnostics) are well recognized by the international APS research community and are utilized by the largest international APS clinical research network registry, APS ACTION, in its core laboratories as the “gold standard” for aPL testing (*86, 87*). Here, IgG, IgM, and IgA aCL assays were reported in GPL, MPL, and APL units, respectively; aβ_2_GPI assays were reported in SGU, SMU, and SAU units; aPS assay were reported in GPS and MPS units; and aPS/PT assays were reported in IgG and IgM units, all per the manufacturer’s specifications. These various units are in accordance with the international consensus guidelines on aPL testing from the 13^th^ International Congress on Antiphospholipid Antibodies (*88*). Per the manufacturer, the establishment of cut-off values for all Quanta Lite® aPL assays are based on balancing sensitivity and specificity to achieve optimal clinical utility. For example, in the case of aPS/PT IgG/IgM (per the manufacturer’s documentation), a total of 91 APS patients, 247 healthy controls, and 43 diseased controls were tested. The threshold chosen resulted in a specificity of 99% for aPS/PT IgG and 98.7% for aPS/PT IgM. A previously described Antiphospholipid Score (**aPL-S**) was used to integrate summarize aPL profiles, with some adaptations(*37*). Here, aPL-S was calculated for each patient by adding points corresponding to the different type and titers of aPL, weighted as below: high-titer aCL IgG (≥40 GPL) = 20 points; low-titer aCL IgG (≥20 GPL) = 4 points; aCL IgM (≥20 MPL) = 2 points; high-titer aβ_2_PGI IgG (≥40 SGU) = 20 points; low-titer aβ_2_PGI IgG (≥20 SGU) = 6 points; aβ_2_PGI IgM (≥20 SMU) = 1 point; high-titer aPS/PT IgG (≥40 units) = 20 points; low-titer aPS/PT IgG (≥30 units) = 13 points; aPS/PT IgM (≥30 units) = 8 points.

### Quantification of S100A8/A9 (calprotectin)

Calprotectin levels were measured with the Human S100A8/S100A9 Heterodimer DuoSet ELISA (DY8226-05, R&D Systems) according to the manufacturer’s instructions.

### Quantification of myeloperoxidase-DNA complexes

Myeloperoxidase-DNA complexes were quantified similarly to what has been previously described(89). This protocol used several reagents from the Cell Death Detection ELISA kit (Roche). First, a high-binding EIA/RIA 96-well plate (Costar) was coated overnight at 4°C with anti-human myeloperoxidase antibody (Bio-Rad 0400-0002), diluted to a concentration of 1 μg/ml in coating buffer (Cell Death kit). The plate was washed two times with wash buffer (0.05% Tween 20 in PBS), and then blocked with 4% bovine serum albumin in PBS (supplemented with 0.05% Tween 20) for 2 hours at room temperature. The plate was again washed five times, before incubating for 90 minutes at room temperature with 10% serum or plasma in the aforementioned blocking buffer (without Tween 20). The plate was washed five times, and then incubated for 90 minutes at room temperature with 10x anti-DNA antibody (HRP-conjugated; from the Cell Death kit) diluted 1:100 in blocking buffer. After five more washes, the plate was developed with 3,3’,5,5’-Tetramethylbenzidine (TMB) substrate (Invitrogen) followed by a 2N sulfuric acid stop solution. Absorbance was measured at a wavelength of 450 nm using a Cytation 5 Cell Imaging Multi-Mode Reader (BioTek). Data were normalized to *in vitro*-prepared NET standards included on every plate, which were quantified based on their DNA content.

### Purification of human IgG

IgG was purified from COVID-19, APS, or control sera with a Protein G Agarose Kit following the manufacturer’s instructions (Pierce). Briefly, serum was diluted in IgG binding buffer and passed through a Protein G Agarose column at least 5 times. IgG was then eluted with 0.1 M glycine and then neutralized with 1 M Tris. This was followed by overnight dialysis against PBS at 4°C. IgG purity was verified with Coomassie staining, and concentrations were determined by BCA protein assay (Pierce) according to manufacturer’s instructions. All IgG samples were determined to have endotoxin level below 0.1 EU/ml by the Pierce™ LAL Chromogenic Endotoxin Quantitation Kit (A39552) according to manufacturer’s instructions. This Kit offers high sensitivity with linear detection range of 0.01-1.0 EU/mL.

### Human neutrophil purification and NETosis assays

Collection of healthy human blood was approved by the University of Michigan IRB (HUM00044257). For neutrophil preparation, blood from healthy volunteers was collected into heparin tubes by standard phlebotomy techniques. The anticoagulated blood was then fractionated by density-gradient centrifugation using Ficoll-Paque Plus (GE Healthcare). Neutrophils were further purified by dextran sedimentation of the red blood cell layer, before lysing residual red blood cells with 0.2% sodium chloride. Neutrophil preparations were at least 95% pure as confirmed by both flow cytometry and nuclear morphology. To assess NETosis, complementary approaches were utilized. For the NET-associated MPO assay, neutrophils were resuspended in RPMI media (Gibco) supplemented with 0.5% bovine serum albumin (BSA, Sigma) and 0.5% fetal bovine serum (Gibco), which had been heat-inactivated at 56°C. Neutrophils (1×10^5^/well) were then incubated in 96-well plates with 10 μg/ml human IgG for 3 hours. To collect NET-associated MPO, the culture media was discarded (to remove any soluble MPO) and replaced with 100 μL of RPMI supplemented with 5 U/ml Micrococcal nuclease (Thermo Fischer Scientific). After 10 minutes at 37°C, digestion of NETs was stopped with 10 mM EDTA. Supernatants were transferred to a v-shaped 96 well plate, and centrifuged at 350xg for 5 minutes to remove debris. Supernatants were then transferred into a new plate. To measure MPO activity, an equal volume of 3,3’,5,5’- Tetramethylbenzidine (TMB) substrate (1 mg ml^-1^, Thermo Fischer Scientific) was added to each well. After 10 minutes of incubation in the dark, the reaction was stopped by the addition of 50 μL of 1 mM sulfuric acid. Absorbance was measured at 450 nm using a Cytation 5 Cell Imaging Multi-Mode Reader. For immunofluorescence microscopy 1.5 × 10^5^ neutrophils were seeded onto coverslips coated with 0.001% poly-L-lysine (Sigma) and fixed with 4% paraformaldehyde. In some experiments, cells were then permeabilized with 0.1% Triton-X for 15 minutes at room temperature. Blocking was with 1% bovine serum albumin. The primary antibody was against neutrophil elastase (Abcam 21595, diluted 1:100), and the FITC-conjugated secondary antibody was from SouthernBiotech (4052-02, diluted 1:250). DNA was stained with Hoechst 33342 (Invitrogen). Images were collected with a Cytation 5 Cell Imaging Multi-Mode Reader.

### Animals and housing

Mice were housed in a specific pathogen-free barrier facility, and fed standard chow. Experimental protocols were approved by the University of Michigan Institutional Animal Care and Use Committee (PRO00008113), and all relevant ethical regulations were followed. Male C57BL/6 mice were purchased from The Jackson Laboratory and used for experiments at 10-12 weeks of age.

### *In vivo* venous thrombosis

To model large-vein thrombosis, we employed procedures that we have utilized previously(*38, 40, 90*). For the stenosis model, a laparotomy was performed under anesthesia. After exposure of the IVC, any lateral branches were ligated using 7-0 Prolene suture (back branches remained patent). A ligature was then fastened around the IVC over a blunted 30-gauge needle (which served as a spacer). After removal of the spacer, the abdomen was closed. Before recovery from anesthesia, mice received a single intravenous injection of human IgG (500 μg). 24 hours later, mice were humanely euthanized, blood was collected, and thrombus characteristics were measured. The electrolytic model was performed as described(*91*). Briefly, after exposure of the IVC, any lateral branches were ligated using 7-0 Prolene suture (back branches remained patent). A 30-gauge silver-coated copper wire (KY-30-1-GRN, Electrospec) with exposed copper wire at the end was placed inside a 25-gauge needle, which was inserted into the IVC and positioned against the anterior wall (where it functioned as the anode). Another needle was implanted subcutaneously, completing the circuit (cathode). A constant current of 250 μA was applied for 15 minutes. The current was supplied by the voltage-to-current converter that is described in detail in the reference(*91*). After removal of the needle, the abdomen was closed. Before recovery from anesthesia, mice received a single intravenous injection of human IgG (500 μg). 24 hours later, mice were humanely euthanized, blood was collected, and thrombus characteristics were measured.

### Western blotting

Thrombi were homogenized in RIPA buffer with Roche protease inhibitor cocktail pellet and 1% SDS. Protein was quantified using the BCA protein assay kit (Pierce). 30 μg of protein was resolved by SDS-PAGE and then transferred to a PVDF membrane. Non-specific binding was blocked with 4% non-fat milk, followed by incubation with primary antibody directed against citrullinated histone H3 (Abcam 5103). Detection was with an HRP-labeled anti-rabbit secondary antibody and an HRP-labeled β-actin antibody, followed by detection using chemiluminescence.

### Thrombin generation assay

Thrombin generation assays were performed using a previously described method (92).

### Statistical analysis

Normally-distributed data were analyzed by two-sided t test and skewed data were analyzed by Mann-Whitney test. Comparisons of more than two groups were analyzed by one-way ANOVA with correction for multiple comparisons by Dunnett’s method. Data analysis was with GraphPad Prism software version 8. Correlations were tested by Spearman’s correlation coefficient. Statistical significance was defined as p<0.05 unless stated otherwise.

## Data Availability

There are no large datasets to deposit. Other data will be made available via correspondence with the authors.

## ACKNOWLEDGEMENTS

The work was supported by a grant from the Burroughs Wellcome Fund to JSK and grants from the Michigan Medicine Frankel Cardiovascular Center and A. Alfred Taubman Medical Research Institute to YK and JSK. YZ was supported by a career development grant from the Rheumatology Research Foundation. JAM was partially supported by the VA Healthcare System. JHM and SAS were supported by the NIH (R35 HL135823). YK was supported by the Intramural Research Program of the NIH and NHLBI, Lasker Foundation, NIH (K08HL131993, R01HL150392), Falk Medical Research Trust Catalyst Award, and the JOBST-American Venous Forum Award. JSK was supported by grants from the NIH (R01HL115138) and Lupus Research Alliance.

## AUTHORSHIP

YZ, SKE, RAA, AAG, SY, HS, GS, KG, JAM, MZ, VY, JW, WW, SPL, and SAS conducted experiments and analyzed data. YZ, NLL, SAS, JHM, YK, and JSK conceived the study and analyzed data. All authors participated in writing the manuscript and gave approval before submission.

## Supplementary Materials

**Supplementary Table 1:** Demographic and clinical characteristics of COVID-19 patients.

|  |  |  |
| --- | --- | --- |
| **Demographics** |  |  |
| Number | 172 |  |
| Age (years)* | 61 ± 17 | (25-95) |
| Female | 75 | (44%) |
| White/Caucasian | 73 | (42%) |
| Black/African-American | 77 | (45%) |
| **Comorbidities** |  |  |
| Diabetes | 70 | (41%) |
| Heart disease | 53 | (30%) |
| Renal disease | 51 | (30%) |
| Lung disease | 47 | (27%) |
| Autoimmune | 9 | (5%) |
| Cancer | 23 | (13%) |
| History of stroke | 16 | (9%) |
| Obesity | 87 | (51%) |
| Hypertension | 117 | (68%) |
| Immune deficiency | 12 | (7%) |
| History of smoking | 45 | (26%) |
| **Medications♦** |  |  |
| Hydroxychloroquine | 31 | (18%) |
| Anti-IL6 receptor | 21 | (12%) |
| ACE inhibitor | 9 | (5%) |
| Angiotensin receptor blocker | 1 | (0.6%) |
| Antibiotic | 66 | (38%) |
| Antiviral | 7 | (4%) |
| **In-hospital thrombosis** |  |  |
| Arterial thrombosis | 2 | (1%) |
| Venous thrombosis | 8 | (5%) |
| Both | 1 | (0.6%) |
| **Final outcomes** |  |  |
| Discharged | 126 | (73%) |
| Death | 33 | (19%) |
| Remains hospitalized | 13 | (8%) |
| * Mean ± standard deviation (range) |  |  |
| ♦ At time of sample collection |  |  |

**Supplementary Table 2:** Prevalence of antiphospholipid antibodies in COVID-19 patients (n=172) based on the first-available sample only.

| aPL | Number positive<br>(manufacturer's cut-off) | % | Number positive<br>(titer ≥40) | % |
| --- | --- | --- | --- | --- |
| aCL IgG | 6 | 3.5% | 1 | 0.58% |
| aCL IgM | 37 | 22% | 11 | 6.4% |
| aCL IgA | 6 | 3.5% | 1 | 0.58% |
| aβ<sub>2</sub>GPI IgG | 5 | 2.9% | 3 | 1.7% |
| aβ<sub>2</sub>GPI IgM | 9 | 5.2% | 7 | 4.1% |
| aβ<sub>2</sub>GPI IgA | 7 | 4.1% | 3 | 1.7% |
| aPS/PT IgG | 37 | 22% | 19 | 11% |
| aPS/PT IgM | 27 | 16% | 19 | 11% |
| any positive aPL | 84 | 49% | 49 | 28% |

aPL=antiphospholipid antibodies; aCL= anticardiolipin; aβ<sub>2</sub>GPI=anti-beta-2 glycoprotein I; aPS/PT=anti-phosphatidylserine/prothrombin Manufacturer's thresholds:
- aCL IgG/M/A=20 GPL/MPL/APL - aβ<sub>2</sub>GPI IgG/M/A=20 SGU/SMU/SAU - aPS/PT IgG/M=30 units

**Supplementary Figure 1:**
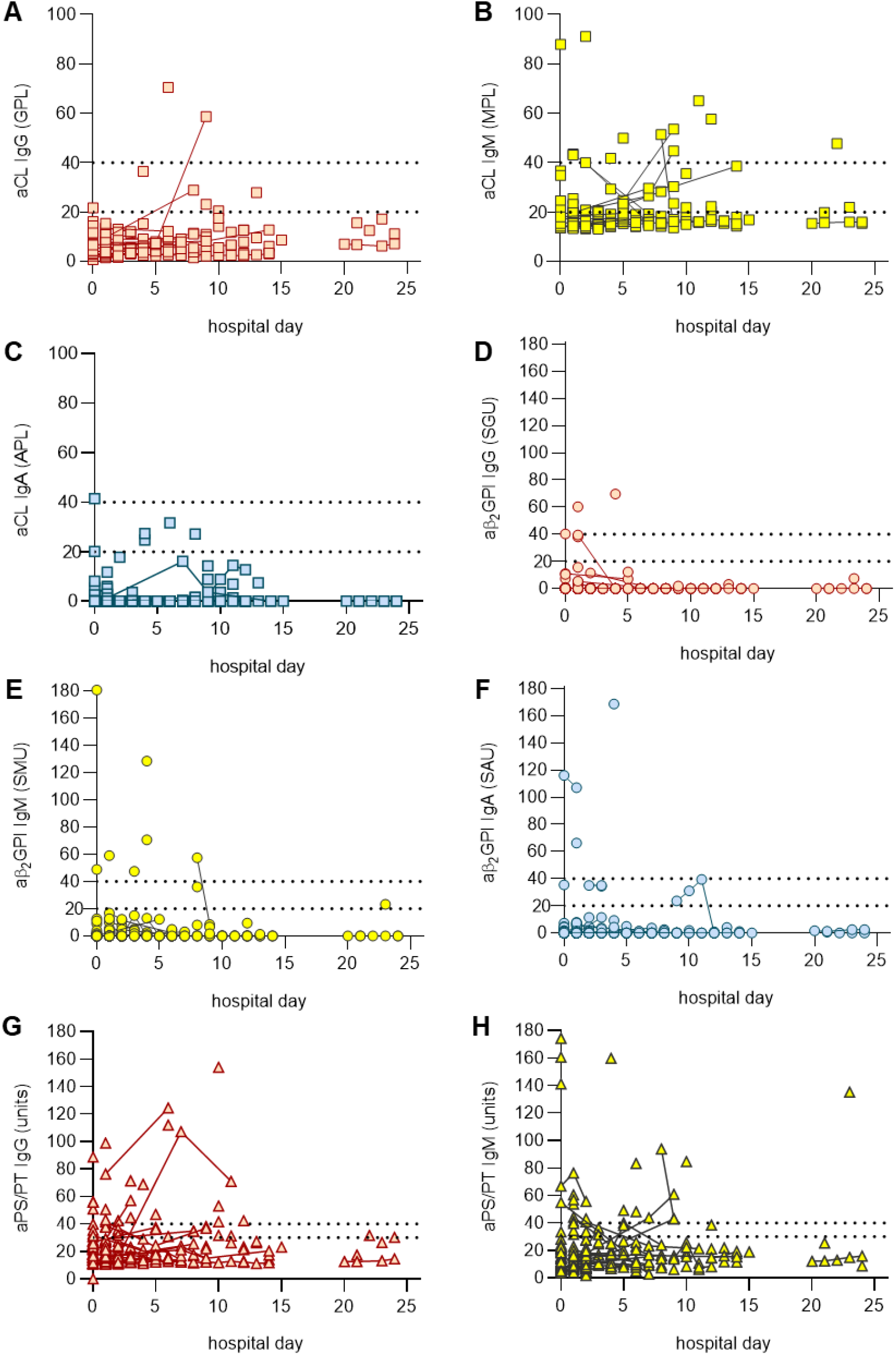
Testing for various aPL presented by day of hospitalization. Repeat testings for the same patient are indicated by solid connecting lines.

**Supplementary Figure 2:**
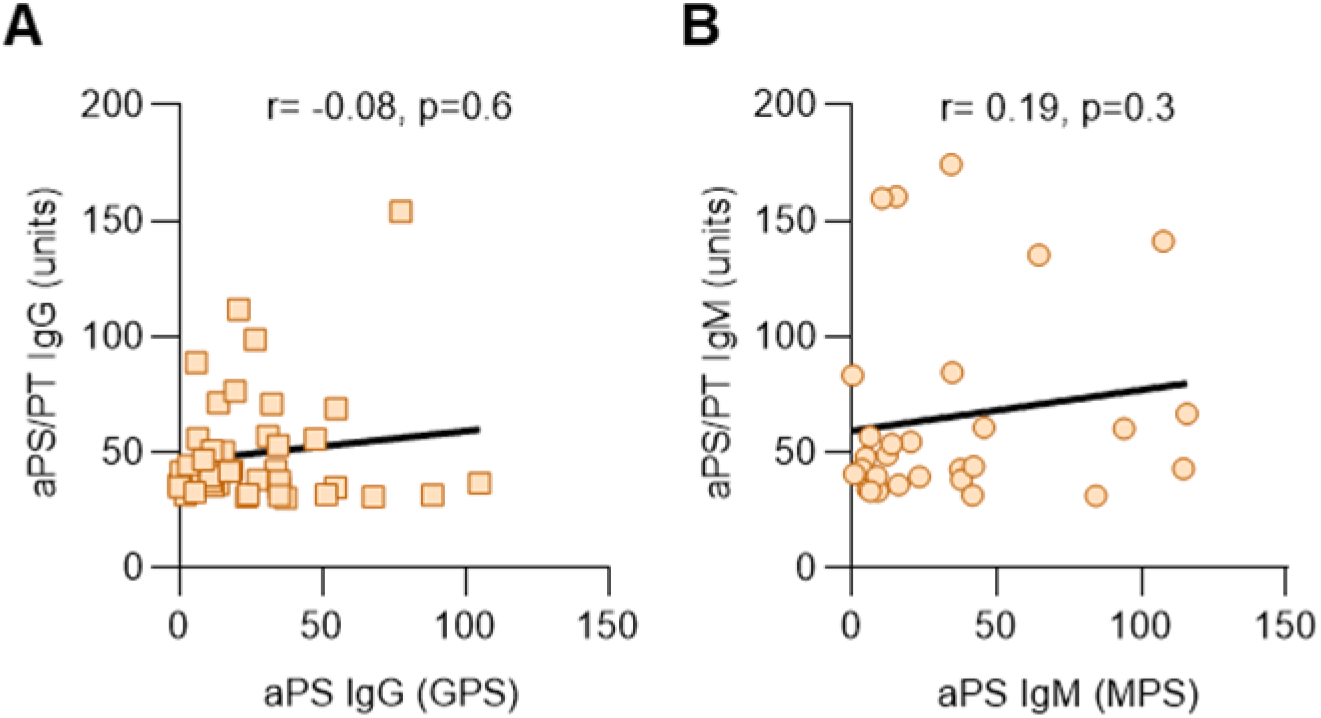
Lack of association between aPS/PT antibodies and aPS antibodies in COVID-19. **A**, Patients with positive aPS/PT IgG were evaluated for aPS IgG. **B**, Patients with positive aPS/PT IgM were evaluated for aPS IgM. Correlations were determined by Spearman’s test. aPS=anti-phosphatidylserine.

**Supplementary Figure 3:**
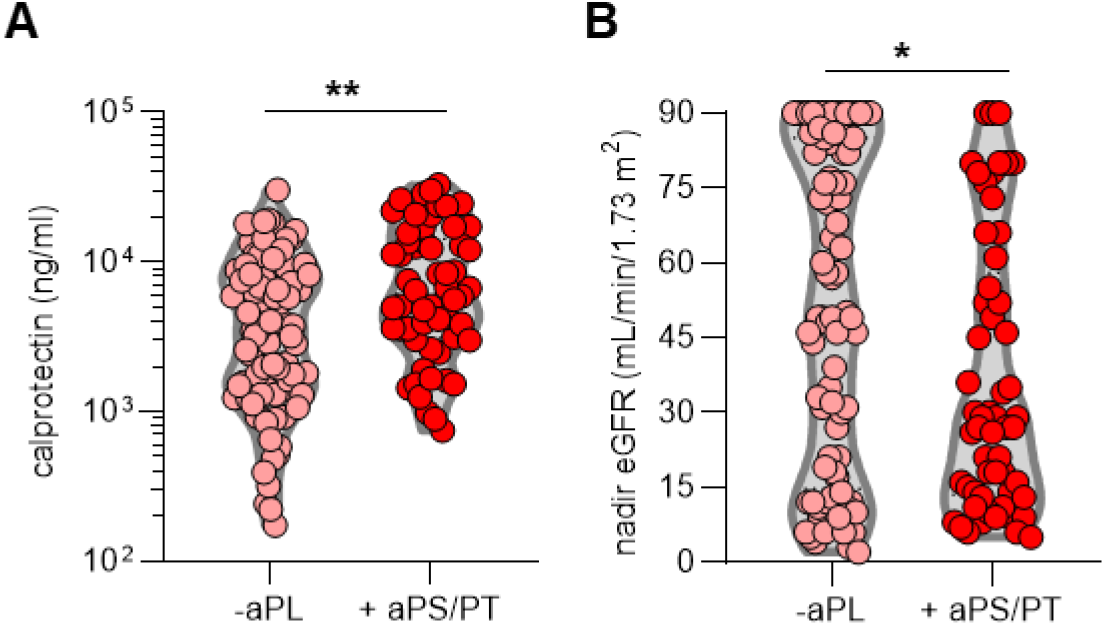
Positive testing for aPL is associated with more neutrophil activation and worse kidney function. **A-B**, COVID-19 patients with positive testing for antiPS/PT (IgG and IgM grouped together) were compared to COVID-19 patients with negative aPL. Manufacturer’s thresholds were used to determine positivity. Groups were analyzed by Mann-Whitney test; *p<0.05, **p<0.01. eGFR=estimated glomerular filtration rate.

**Supplementary Figure 4:**
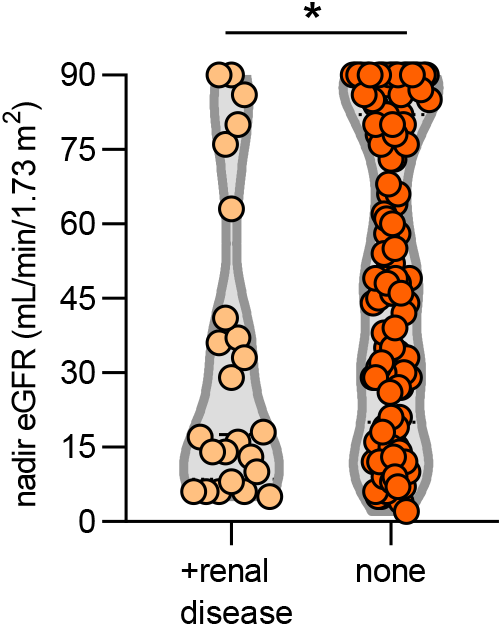
Nadir eGFR in COVID-19 patients with and without prior history of renal disease. Groups were analyzed by Mann-Whitney test; *p<0.05. eGFR=estimated glomerular filtration rate.

**Supplementary Figure 5:**
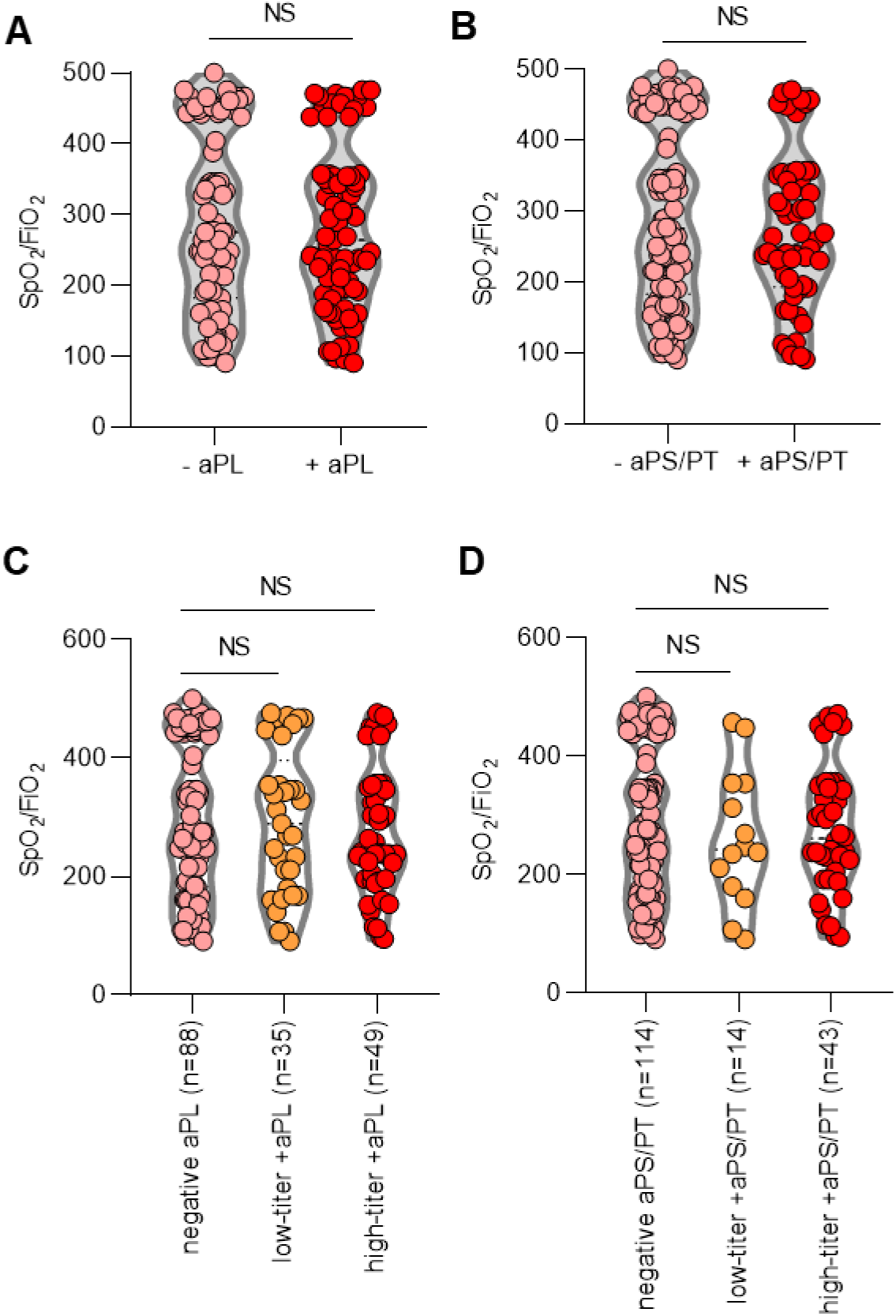
aPL status as a predictor of oxygenation efficiency. **A-B**, COVID-19 patients were divided into groups based on positivity (manufacturer’s thresholds) for any aPL (A) or aPS/PT (B). **C**, Patients were placed into groups according to high-titer positive aPL (>40 units), low-titer positive aPL (20-40 units), or negative for any aPL. **D**, Patients were placed into groups according to high-titer positive aPS/PT (>40 units), low-titer positive aPS/PT (30-40 units), or negative for any aPL. Oxygenation efficiency (SpO_2_/FiO_2_) was compared by Mann-Whitney test (A-B) or Kruskal-Wallis test (C-D); NS=not significant.

**Supplementary Figure 6:**
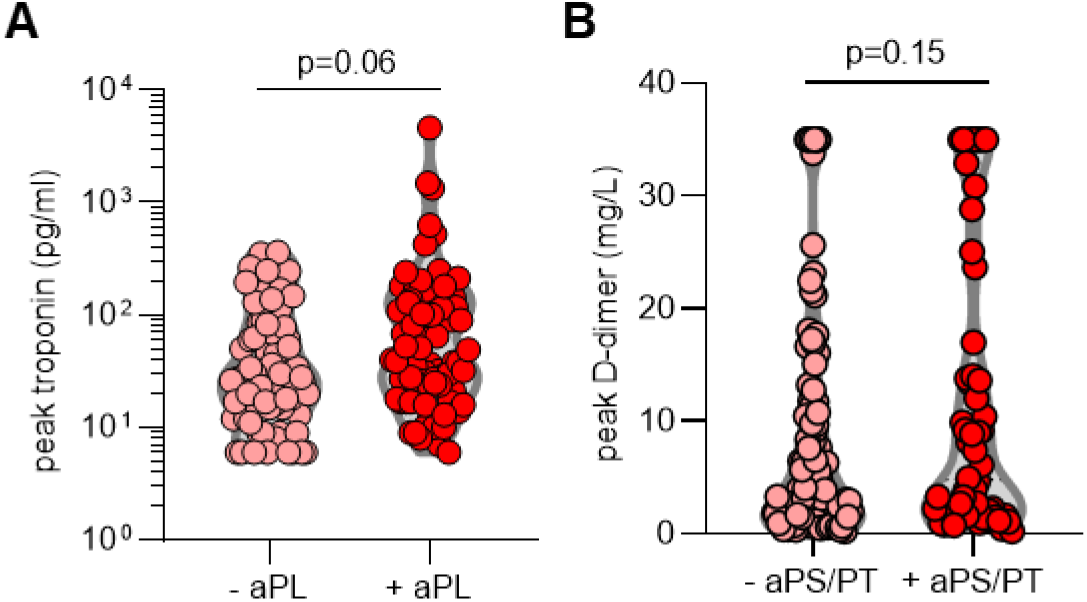
aPL status as a predictor of peak troponin and D-dimer. **A**, COVID-19 patients were divided into groups based on positivity (manufacturer’s thresholds) for any aPL. **B**, COVID-19 patients were grouped based on positive testing for aPS/PT (IgG and IgM grouped together); manufacturer’s thresholds were used to determine positivity. Groups were compared by unpaired t-test.

**Supplementary Figure 7:**
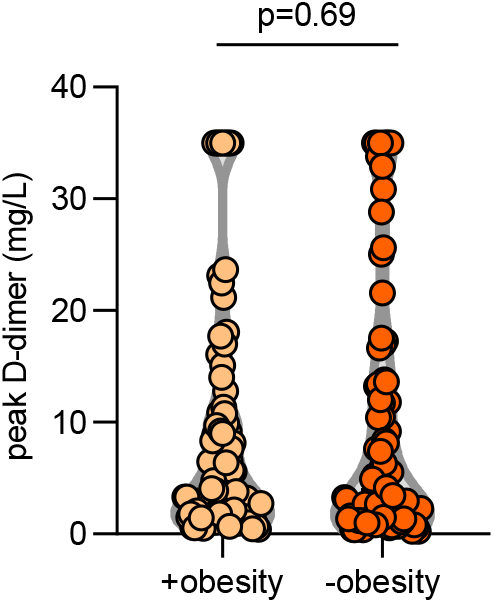
Levels of D-dimer in patients with and without obesity. Obesity was defined as body mass index of 30 or greater. Groups were compared by Mann-Whitney test.

**Supplementary Figure 8:**
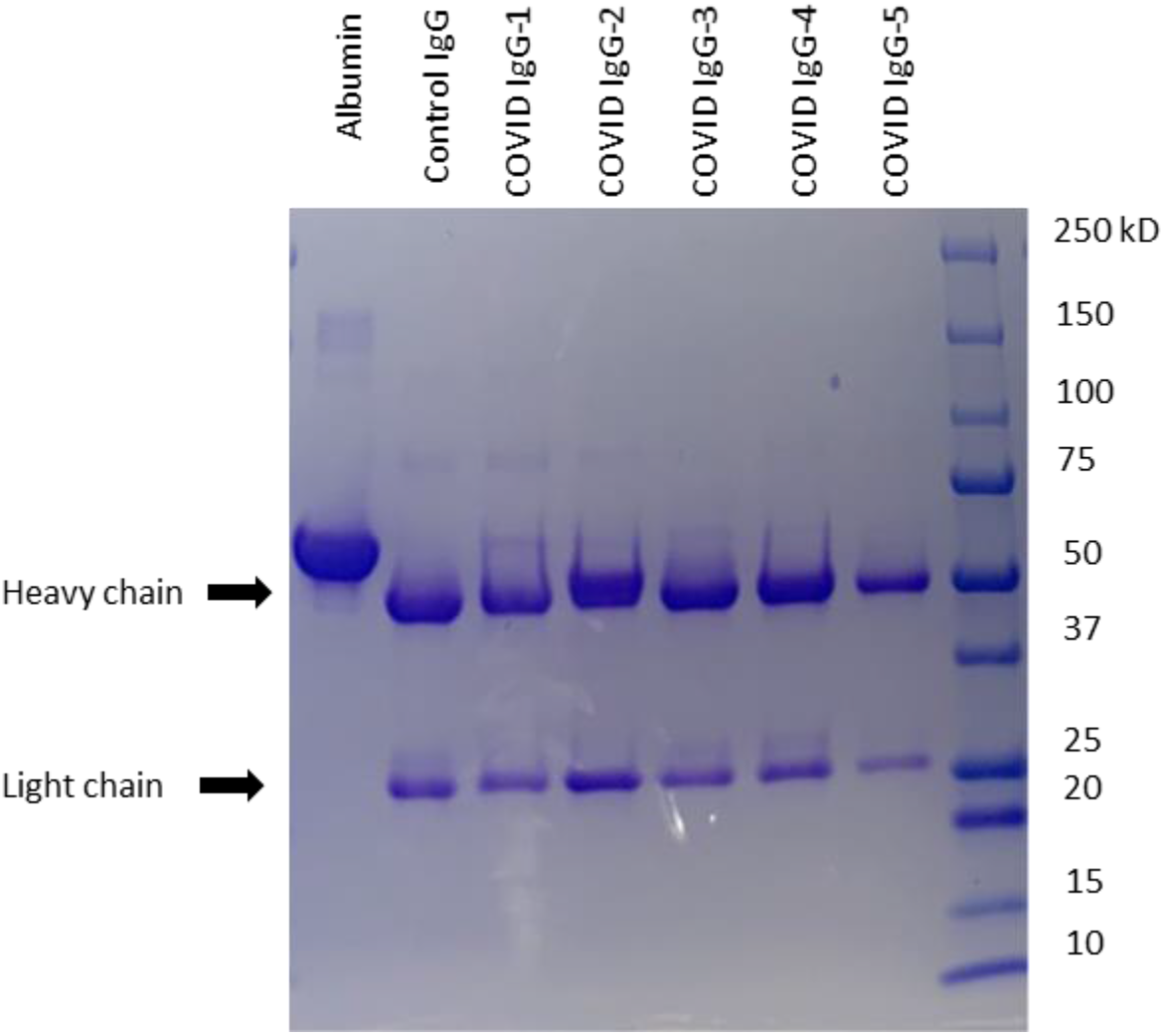
Determination of COVID-19 IgG purity. COVID-19 IgG was resolved by SDS-PAGE and stained by Coomassie Brilliant Blue.

**Supplementary Figure 9:**
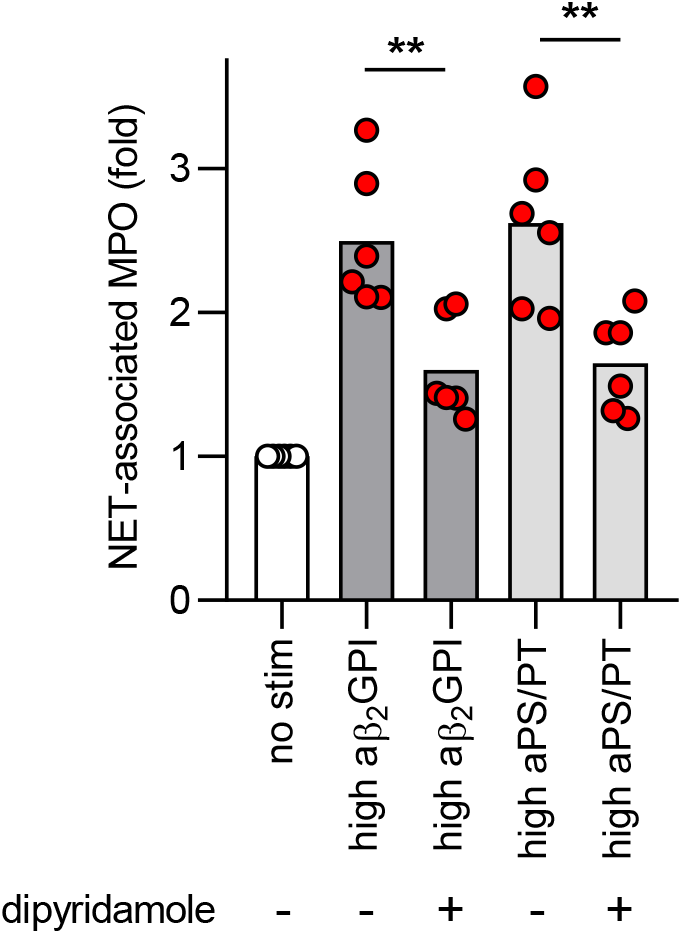
Dipyridamole suppresses COVID IgG-induced neutrophil extracellular trap (NET) release. Neutrophils were isolated from healthy controls and cultured with COVID-19 IgG (10 μg/ml) for 3 hours. Some samples were additionally treated with 10 μM dipyridamole. NET release was measured by the enzymatic activity of myeloperoxidase (MPO) after solubilization of NETs with Micrococcal nuclease. Data are presented for four independent experiments. Comparisons were by one-way ANOVA with correction for multiple comparisons by Dunnett’s method; **p<0.01.

**Supplementary Figure 10:**
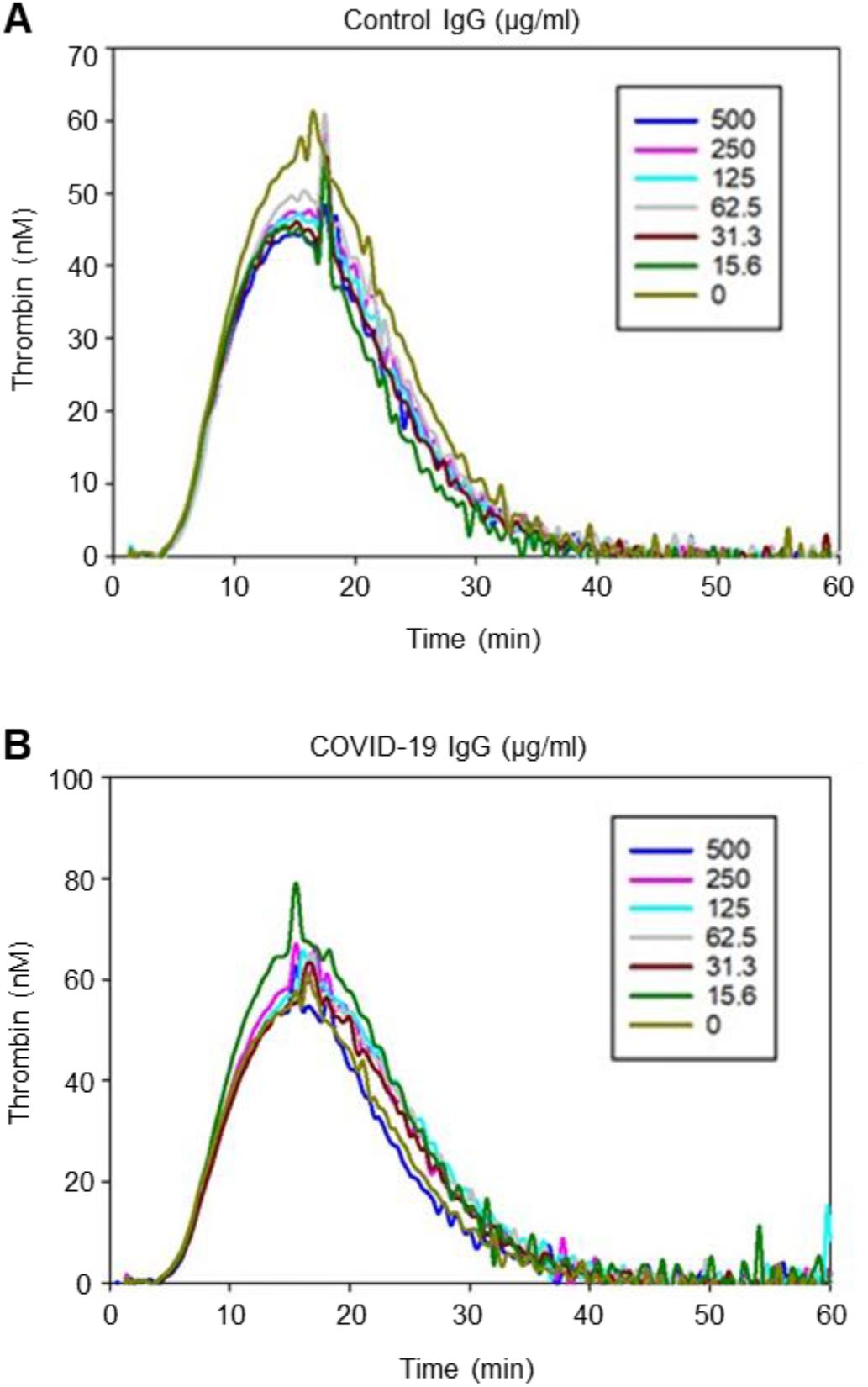
COVID-19 IgG does not potentiate thrombin generation in cell-free pooled normal plasma. Preparations of control IgG or COVID-19 IgG (a high-aPS/PT sample) were titrated into pooled normal plasma at multiple concentrations as indicated. Thrombin generation was triggered by the addition of 1 pM tissue factor. These data are representative of three independent experiments.

**Supplementary Figure 11:**
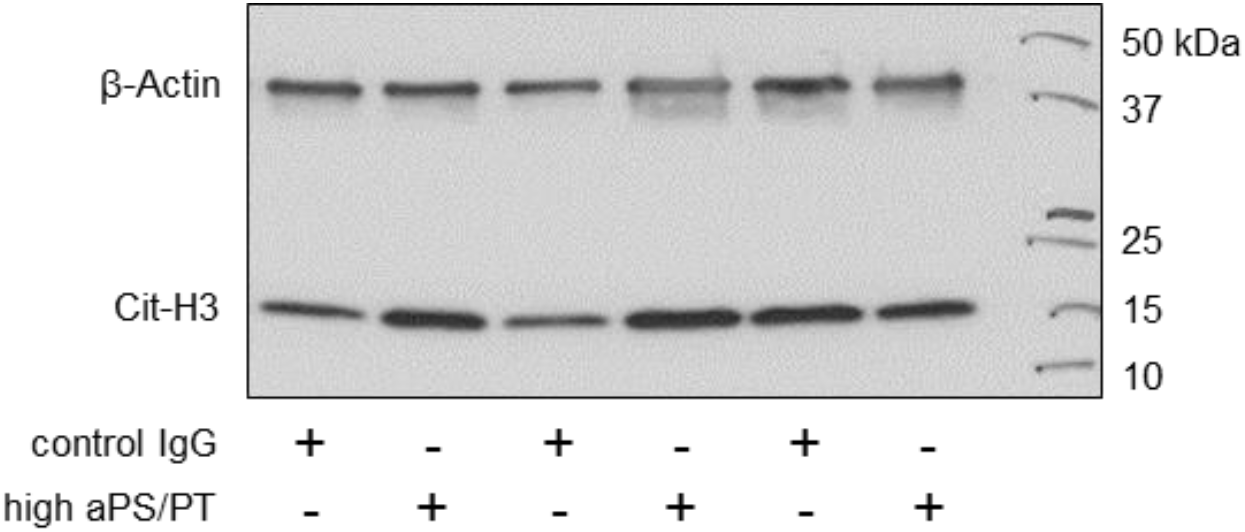
Measurement of citrullinated histone H3 (Cit-H3) in mouse thrombi. Mice were treated with control IgG or high-aPS/PT COVID-19 IgG as for Figure 3. Inferior vena cava thrombi were triggered by electrolytic free radicals. After 24 hours, thrombi were isolated and total thrombus protein was resolved by SDS-PAGE; both beta-actin and Cit-H3 were detected by western blotting.

## REFERENCES

1. C. Wu, X. Chen, Y. Cai, J. Xia, X. Zhou, S. Xu, H. Huang, L. Zhang, X. Zhou, C. Du, Y. Zhang, J. Song, S. Wang, Y. Chao, Z. Yang, J. Xu, X. Zhou, D. Chen, W. Xiong, L. Xu, F. Zhou, J. Jiang, C. Bai, J. Zheng, Y. Song, Risk Factors Associated With Acute Respiratory Distress Syndrome and Death in Patients With Coronavirus Disease 2019 Pneumonia in Wuhan, China. JAMA Intern Med, (2020).

2. N. Tang, D. Li, X. Wang, Z. Sun, Abnormal coagulation parameters are associated with poor prognosis in patients with novel coronavirus pneumonia. J Thromb Haemost 18, 844–847 (2020).

3. W. J. Guan, Z. Y. Ni, Y. Hu, W. H. Liang, C. Q. Ou, J. X. He, L. Liu, H. Shan, C. L. Lei, D. S. C. Hui, B. Du, L. J. Li, G. Zeng, K. Y. Yuen, R. C. Chen, C. L. Tang, T. Wang, P. Y. Chen, J. Xiang, S. Y. Li, J. L. Wang, Z. J. Liang, Y. X. Peng, L. Wei, Y. Liu, Y. H. Hu, P. Peng, J. M. Wang, J. Y. Liu, Z. Chen, G. Li, Z. J. Zheng, S. Q. Qiu, J. Luo, C. J. Ye, S. Y. Zhu, N. S. Zhong, C. China Medical Treatment Expert Group for, Clinical Characteristics of Coronavirus Disease 2019 in China. N Engl J Med, (2020).

4. N. Chen, M. Zhou, X. Dong, J. Qu, F. Gong, Y. Han, Y. Qiu, J. Wang, Y. Liu, Y. Wei, J. Xia, T. Yu, X. Zhang, L. Zhang, Epidemiological and clinical characteristics of 99 cases of 2019 novel coronavirus pneumonia in Wuhan, China: a descriptive study. Lancet 395, 507–513 (2020).

5. F. Zhou, T. Yu, R. Du, G. Fan, Y. Liu, Z. Liu, J. Xiang, Y. Wang, B. Song, X. Gu, L. Guan, Y. Wei, H. Li, X. Wu, J. Xu, S. Tu, Y. Zhang, H. Chen, B. Cao, Clinical course and risk factors for mortality of adult inpatients with COVID-19 in Wuhan, China: a retrospective cohort study. Lancet 395, 1054–1062 (2020).

6. T. Iba, J. H. Levy, T. E. Warkentin, J. Thachil, T. van der Poll, M. Levi, Scientific, D. I. C. Standardization Committee on, S. the, P. Standardization Committee on, T. Critical Care of the International Society on, Haemostasis, Diagnosis and management of sepsis-induced coagulopathy and disseminated intravascular coagulation. J Thromb Haemost 17, 1989–1994 (2019).

7. T. Iba, M. Di Nisio, J. Thachil, H. Wada, H. Asakura, K. Sato, D. Saitoh, A Proposal of the Modification of Japanese Society on Thrombosis and Hemostasis (JSTH) Disseminated Intravascular Coagulation (DIC) Diagnostic Criteria for Sepsis-Associated DIC. Clin Appl Thromb Hemost 24, 439–445 (2018).

8. S. Cui, S. Chen, X. Li, S. Liu, F. Wang, Prevalence of venous thromboembolism in patients with severe novel coronavirus pneumonia. J Thromb Haemost, (2020).

9. F. A. Klok, M. Kruip, N. J. M. van der Meer, M. S. Arbous, D. Gommers, K. M. Kant, F. H. J. Kaptein, J. van Paassen, M. A. M. Stals, M. V. Huisman, H. Endeman, Incidence of thrombotic complications in critically ill ICU patients with COVID-19. Thromb Res, (2020).

10. C. Lodigiani, G. Iapichino, L. Carenzo, M. Cecconi, P. Ferrazzi, T. Sebastian, N. Kucher, J. D. Studt, C. Sacco, B. Alexia, M. T. Sandri, S. Barco, C.-T. F. Humanitas, Venous and arterial thromboembolic complications in COVID-19 patients admitted to an academic hospital in Milan, Italy. Thromb Res 191, 9–14 (2020).

11. S. Tian, W. Hu, L. Niu, H. Liu, H. Xu, S. Y. Xiao, Pulmonary Pathology of Early-Phase 2019 Novel Coronavirus (COVID-19) Pneumonia in Two Patients With Lung Cancer. J Thorac Oncol, (2020).

12. X. Liu, Z. Li, S. Liu, Z. Chen, Z. Zhao, Y.-y. Huang, Q. Zhang, J. Wang, Y. Shi, Y. Xu, J. Sun, H. Xian, R. Fang, F. Bai, C. Ou, B. Xiong, A. M. Lew, J. Cui, H. Huang, J. Zhao, X. Hong, Y. Zhang, F. Zhou, H.-B. Luo, Therapeutic effects of dipyridamole on COVID-19 patients with coagulation dysfunction. 2020.2002.2027.20027557 (2020).

13. S. E. Fox, A. Akmatbekov, J. L. Harbert, G. Li, J. Quincy Brown, R. S. Vander Heide, Pulmonary and cardiac pathology in African American patients with COVID-19: an autopsy series from New Orleans. Lancet Respir Med, (2020).

14. M. E. Colling, Y. Kanthi, COVID-19-associated coagulopathy: An exploration of mechanisms. Vasc Med, 1358863X20932640 (2020).

15. Y. Zuo, S. Yalavarthi, H. Shi, K. Gockman, M. Zuo, J. A. Madison, C. Blair, A. Weber, B. J. Barnes, M. Egeblad, R. J. Woods, Y. Kanthi, J. S. Knight, Neutrophil extracellular traps in COVID-19. JCI Insight 5, (2020).

16. H. Shi, Y. Zuo, S. Yalavarthi, K. Gockman, M. Zuo, J. A. Madison, C. Blair, W. Woodward, S. P. Lezak, N. L. Lugogo, R. J. Woods, C. Lood, J. S. Knight, Y. Kanthi, Neutrophil calprotectin identifies severe pulmonary disease in COVID-19. J Leukoc Biol, (2020).

17. B. J. Barnes, J. M. Adrover, A. Baxter-Stoltzfus, A. Borczuk, J. Cools-Lartigue, J. M. Crawford, J. Dassler-Plenker, P. Guerci, C. Huynh, J. S. Knight, M. Loda, M. R. Looney, F. McAllister, R. Rayes, S. Renaud, S. Rousseau, S. Salvatore, R. E. Schwartz, J. D. Spicer, C. C. Yost, A. Weber, Y. Zuo, M. Egeblad, Targeting potential drivers of COVID-19: Neutrophil extracellular traps. J Exp Med 217, (2020).

18. M. Leppkes, J. Knopf, E. Naschberger, A. Lindemann, J. Singh, I. Herrmann, M. Sturzl, L. Staats, A. Mahajan, C. Schauer, A. N. Kremer, S. Volkl, K. Amann, K. Evert, C. Falkeis, A. Wehrfritz, R. J. Rieker, A. Hartmann, A. E. Kremer, M. F. Neurath, L. E. Munoz, G. Schett, M. Herrmann, Vascular occlusion by neutrophil extracellular traps in COVID-19. EBioMedicine 58, 102925 (2020).

19. E. A. Middleton, X. Y. He, F. Denorme, R. A. Campbell, D. Ng, S. P. Salvatore, M. Mostyka, A. Baxter-Stoltzfus, A. C. Borczuk, M. Loda, M. J. Cody, B. K. Manne, I. Portier, E. S. Harris, A. C. Petrey, E. J. Beswick, A. F. Caulin, A. Iovino, L. M. Abegglen, A. S. Weyrich, M. T. Rondina, M. Egeblad, J. D. Schiffman, C. C. Yost, Neutrophil extracellular traps contribute to immunothrombosis in COVID-19 acute respiratory distress syndrome. Blood 136, 1169–1179 (2020).

20. L. Nicolai, A. Leunig, S. Brambs, R. Kaiser, T. Weinberger, M. Weigand, M. Muenchhoff, J. C. Hellmuth, S. Ledderose, H. Schulz, C. Scherer, M. Rudelius, M. Zoller, D. Hochter, O. Keppler, D. Teupser, B. Zwissler, M. Bergwelt-Baildon, S. Kaab, S. Massberg, K. Pekayvaz, K. Stark, Immunothrombotic Dysregulation in COVID-19 Pneumonia is Associated with Respiratory Failure and Coagulopathy. Circulation, (2020).

21. J. A. Madison, A. Duarte-Garcia, Y. Zuo, J. S. Knight, Treatment of thrombotic antiphospholipid syndrome in adults and children. Curr Opin Rheumatol 32, 215–227 (2020).

22. S. Yalavarthi, T. J. Gould, A. N. Rao, L. F. Mazza, A. E. Morris, C. Nunez-Alvarez, D. Hernandez-Ramirez, P. L. Bockenstedt, P. C. Liaw, A. R. Cabral, J. S. Knight, Release of neutrophil extracellular traps by neutrophils stimulated with antiphospholipid antibodies: a newly identified mechanism of thrombosis in the antiphospholipid syndrome. Arthritis Rheumatol 67, 2990–3003 (2015).

23. Y. Zuo, H. Shi, C. Li, J. S. Knight, Antiphospholipid syndrome: a clinical perspective. Chin Med J (Engl), (2020).

24. N. M. Kazzaz, W. J. McCune, J. S. Knight, Treatment of catastrophic antiphospholipid syndrome. Curr Opin Rheumatol 28, 218–227 (2016).

25. S. Miyakis, M. D. Lockshin, T. Atsumi, D. W. Branch, R. L. Brey, R. Cervera, R. H. Derksen, D. E. G. Pg, T. Koike, P. L. Meroni, G. Reber, Y. Shoenfeld, A. Tincani, P. G. Vlachoyiannopoulos, S. A. Krilis, International consensus statement on an update of the classification criteria for definite antiphospholipid syndrome (APS). J Thromb Haemost 4, 295–306 (2006).

26. H. Shi, H. Zheng, Y. F. Yin, Q. Y. Hu, J. L. Teng, Y. Sun, H. L. Liu, X. B. Cheng, J. N. Ye, Y. T. Su, X. Y. Wu, J. F. Zhou, G. L. Norman, H. Y. Gong, X. M. Shi, Y. B. Peng, X. F. Wang, C. D. Yang, Antiphosphatidylserine/prothrombin antibodies (aPS/PT) as potential diagnostic markers and risk predictors of venous thrombosis and obstetric complications in antiphospholipid syndrome. Clin Chem Lab Med 56, 614–624 (2018).

27. K. M. J. Devreese, E. A. Linskens, D. Benoit, H. Peperstraete, Antiphospholipid antibodies in patients with COVID-19: A relevant observation? J Thromb Haemost, (2020).

28. Y. Zhang, M. Xiao, S. Zhang, P. Xia, W. Cao, W. Jiang, H. Chen, X. Ding, H. Zhao, H. Zhang, C. Wang, J. Zhao, X. Sun, R. Tian, W. Wu, D. Wu, J. Ma, Y. Chen, D. Zhang, J. Xie, X. Yan, X. Zhou, Z. Liu, J. Wang, B. Du, Y. Qin, P. Gao, X. Qin, Y. Xu, W. Zhang, T. Li, F. Zhang, Y. Zhao, Y. Li, S. Zhang, Coagulopathy and Antiphospholipid Antibodies in Patients with Covid-19. N Engl J Med 382, e38 (2020).

29. M. Gatto, C. Perricone, M. Tonello, O. Bistoni, A. M. Cattelan, R. Bursi, G. Cafaro, E. De Robertis, A. Mencacci, S. Bozza, A. Vianello, L. Iaccarino, R. Gerli, A. Doria, E. Bartoloni, Frequency and clinical correlates of antiphospholipid antibodies arising in patients with SARS-CoV-2 infection: findings from a multicentre study on 122 cases. Clin Exp Rheumatol 38, 754–759 (2020).

30. V. Siguret, S. Voicu, M. Neuwirth, M. Delrue, E. Gayat, A. Stepanian, B. Megarbane, Are antiphospholipid antibodies associated with thrombotic complications in critically ill COVID-19 patients? Thromb Res 195, 74–76 (2020).

31. M. Xiao, Y. Zhang, S. Zhang, X. Qin, P. Xia, W. Cao, W. Jiang, H. Chen, X. Ding, H. Zhao, H. Zhang, C. Wang, J. Zhao, X. Sun, R. Tian, W. Wu, D. Wu, J. Ma, Y. Chen, D. Zhang, J. Xie, X. Yan, X. Zhou, Z. Liu, J. Wang, B. Du, Y. Qin, P. Gao, M. Lu, X. Hou, X. Wu, H. Zhu, Y. Xu, W. Zhang, T. Li, F. Zhang, Y. Zhao, Y. Li, S. Zhang, Brief Report: Anti-phospholipid antibodies in critically ill patients with Coronavirus Disease 2019 (COVID-19). Arthritis Rheumatol, (2020).

32. M. O. Borghi, A. Beltagy, E. Garrafa, D. Curreli, G. Cecchini, C. Bodio, C. Grossi, S. Blengino, A. Tincani, F. Franceschini, L. Andreoli, M. G. Lazzaroni, S. Piantoni, S. Masneri, F. Crisafulli, D. Brugnoni, M. L. Muiesan, M. Salvetti, G. Parati, E. Torresani, M. Mahler, F. Heilbron, F. Pregnolato, M. Pengo, F. Tedesco, N. Pozzi, P. L. Meroni, Prevalence, specificity, and clinical association of anti-phospholipid antibodies in COVID-19 patients: are the antibodies really guilty? *medRxiv,* (2020).

33. N. Abdel-Wahab, S. Talathi, M. A. Lopez-Olivo, M. E. Suarez-Almazor, Risk of developing antiphospholipid antibodies following viral infection: a systematic review and meta-analysis. Lupus 27, 572–583 (2018).

34. R. A. Asherson, R. Cervera, ‘Primary’, ‘secondary’ and other variants of the antiphospholipid syndrome. Lupus 3, 293–298 (1994).

35. N. Abdel-Wahab, M. A. Lopez-Olivo, G. P. Pinto-Patarroyo, M. E. Suarez-Almazor, Systematic review of case reports of antiphospholipid syndrome following infection. Lupus 25, 1520–1531 (2016).

36. J. Sung, S. Anjum, Coronavirus Disease 2019 (COVID-19) Infection Associated With Antiphospholipid Antibodies and Four-Extremity Deep Vein thrombosis in a Previously Healthy Female. Cureus 12, e8408 (2020).

37. K. Otomo, T. Atsumi, O. Amengual, Y. Fujieda, M. Kato, K. Oku, T. Horita, S. Yasuda, T. Koike, Efficacy of the antiphospholipid score for the diagnosis of antiphospholipid syndrome and its predictive value for thrombotic events. Arthritis Rheum 64, 504–512 (2012).

38. H. Meng, S. Yalavarthi, Y. Kanthi, L. F. Mazza, M. A. Elfline, C. E. Luke, D. J. Pinsky, P. K. Henke, J. S. Knight, In Vivo Role of Neutrophil Extracellular Traps in Antiphospholipid Antibody-Mediated Venous Thrombosis. Arthritis Rheumatol 69, 655–667 (2017).

39. R. A. Ali, A. A. Gandhi, H. Meng, S. Yalavarthi, A. P. Vreede, S. K. Estes, O. R. Palmer, P. L. Bockenstedt, D. J. Pinsky, J. M. Greve, J. A. Diaz, Y. Kanthi, J. S. Knight, Adenosine receptor agonism protects against NETosis and thrombosis in antiphospholipid syndrome. Nat Commun 10, 1916 (2019).

40. J. S. Knight, H. Meng, P. Coit, S. Yalavarthi, G. Sule, A. A. Gandhi, R. C. Grenn, L. F. Mazza, R. A. Ali, P. Renauer, J. D. Wren, P. L. Bockenstedt, H. Wang, D. T. Eitzman, A. H. Sawalha, Activated signature of antiphospholipid syndrome neutrophils reveals potential therapeutic target. JCI Insight 2, (2017).

41. Y. Zuo, H. Shi, C. Li, J. S. Knight, Antiphospholipid syndrome: a clinical perspective. Chin Med J (Engl) 133, 929–940 (2020).

42. K. L. Allen, F. V. Fonseca, V. Betapudi, B. Willard, J. Zhang, K. R. McCrae, A novel pathway for human endothelial cell activation by antiphospholipid/anti-beta2 glycoprotein I antibodies. Blood 119, 884–893 (2012).

43. M. Galli, P. Comfurius, C. Maassen, H. C. Hemker, M. H. de Baets, P. J. van Breda-Vriesman, T. Barbui, R. F. Zwaal, E. M. Bevers, Anticardiolipin antibodies (ACA) directed not to cardiolipin but to a plasma protein cofactor. Lancet 335, 1544–1547 (1990).

44. O. Amengual, T. Atsumi, T. Koike, Antiprothombin antibodies and the diagnosis of antiphospholipid syndrome. Clin Immunol 112, 144–149 (2004).

45. P. Permpikul, L. V. Rao, S. I. Rapaport, Functional and binding studies of the roles of prothrombin and beta 2-glycoprotein I in the expression of lupus anticoagulant activity. Blood 83, 2878–2892 (1994).

46. P. P. Chen, I. Giles, Antibodies to serine proteases in the antiphospholipid syndrome. Curr Rheumatol Rep 12, 45–52 (2010).

47. G. Cesarman-Maus, N. P. Rios-Luna, A. B. Deora, B. Huang, R. Villa, C. Cravioto Mdel, D. Alarcon-Segovia, J. Sanchez-Guerrero, K. A. Hajjar, Autoantibodies against the fibrinolytic receptor, annexin 2, in antiphospholipid syndrome. Blood 107, 4375–4382 (2006).

48. M. Blank, I. Krause, M. Fridkin, N. Keller, J. Kopolovic, I. Goldberg, A. Tobar, Y. Shoenfeld, Bacterial induction of autoantibodies to beta2-glycoprotein-I accounts for the infectious etiology of antiphospholipid syndrome. J Clin Invest 109, 797–804 (2002).

49. A. E. Gharavi, S. S. Pierangeli, R. G. Espinola, X. Liu, M. Colden-Stanfield, E. N. Harris, Antiphospholipid antibodies induced in mice by immunization with a cytomegalovirus-derived peptide cause thrombosis and activation of endothelial cells in vivo. Arthritis Rheum 46, 545–552 (2002).

50. P. Cruz-Tapias, M. Blank, J. M. Anaya, Y. Shoenfeld, Infections and vaccines in the etiology of antiphospholipid syndrome. Curr Opin Rheumatol 24, 389–393 (2012).

51. R. Cervera, R. A. Asherson, M. L. Acevedo, J. A. Gomez-Puerta, G. Espinosa, G. De La Red, V. Gil, M. Ramos-Casals, M. Garcia-Carrasco, M. Ingelmo, J. Font, Antiphospholipid syndrome associated with infections: clinical and microbiological characteristics of 100 patients. Ann Rheum Dis 63, 1312–1317 (2004).

52. P. G. de Groot, R. T. Urbanus, The significance of autoantibodies against beta2-glycoprotein I. Blood 120, 266–274 (2012).

53. M. L. Durkin, D. Marchese, M. D. Robinson, M. Ramgopal, Catastrophic antiphospholipid syndrome (CAPS) induced by influenza A virus subtype H1N1. BMJ Case Rep 2013, (2013).

54. J. E. Hunt, H. P. McNeil, G. J. Morgan, R. M. Crameri, S. A. Krilis, A phospholipid-beta 2-glycoprotein I complex is an antigen for anticardiolipin antibodies occurring in autoimmune disease but not with infection. Lupus 1, 75–81 (1992).

55. M. Blank, R. A. Asherson, R. Cervera, Y. Shoenfeld, Antiphospholipid syndrome infectious origin. J Clin Immunol 24, 12–23 (2004).

56. M. Yamazaki, H. Asakura, Y. Kawamura, T. Ohka, M. Endo, T. Matsuda, Transient lupus anticoagulant induced by Epstein-Barr virus infection. Blood Coagul Fibrinolysis 2, 771–774 (1991).

57. P. von Landenberg, H. W. Lehmann, S. Modrow, Human parvovirus B19 infection and antiphospholipid antibodies. Autoimmun Rev 6, 278–285 (2007).

58. C. Catoggio, A. Alvarez-Uria, P. L. Fernandez, R. Cervera, G. Espinosa, Catastrophic antiphospholipid syndrome triggered by fulminant disseminated herpes simplex infection in a patient with systemic lupus erythematosus. Lupus 21, 1359–1361 (2012).

59. R. A. Asherson, R. Cervera, Antiphospholipid antibodies and infections. Ann Rheum Dis 62, 388–393 (2003).

60. R. Cervera, I. Rodriguez-Pinto, G. Espinosa, The diagnosis and clinical management of the catastrophic antiphospholipid syndrome: A comprehensive review. J Autoimmun 92, 1–11 (2018).

61. I. Rodriguez-Pinto, M. Moitinho, I. Santacreu, Y. Shoenfeld, D. Erkan, G. Espinosa, R. Cervera, C. R. P. Group, Catastrophic antiphospholipid syndrome (CAPS): Descriptive analysis of 500 patients from the International CAPS Registry. Autoimmun Rev 15, 1120–1124 (2016).

62. S. Kapoor, A. Opneja, L. Nayak, The role of neutrophils in thrombosis. Thromb Res 170, 87–96 (2018).

63. C. Thalin, Y. Hisada, S. Lundstrom, N. Mackman, H. Wallen, Neutrophil Extracellular Traps: Villains and Targets in Arterial, Venous, and Cancer-Associated Thrombosis. Arterioscler Thromb Vasc Biol 39, 1724–1738 (2019).

64. G. Sule, W. J. Kelley, K. Gockman, S. Yalavarthi, A. P. Vreede, A. L. Banka, P. L. Bockenstedt, O. Eniola-Adefeso, J. S. Knight, Increased Adhesive Potential of Antiphospholipid Syndrome Neutrophils Mediated by beta2 Integrin Mac-1. Arthritis Rheumatol 72, 114–124 (2020).

65. Y. Kanthi, J. S. Knight, Y. Zuo, D. J. Pinsky, New (re)Purpose for an old drug: purinergic receptor blockade may extinguish the COVID-19 thrombo-inflammatory firestorm. JCI Insight, (2020).

66. X. Liu, Z. Li, S. Liu, J. Sun, Z. Chen, M. Jiang, Q. Zhang, Y. Wei, X. Wang, Y. Y. Huang, Y. Shi, Y. Xu, H. Xian, F. Bai, C. Ou, B. Xiong, A. M. Lew, J. Cui, R. Fang, H. Huang, J. Zhao, X. Hong, Y. Zhang, F. Zhou, H. B. Luo, Potential therapeutic effects of dipyridamole in the severely ill patients with COVID-19. Acta Pharm Sin B, (2020).

67. Y. Kanthi, J. S. Knight, Y. Zuo, D. J. Pinsky, New (re)purpose for an old drug: purinergic modulation may extinguish the COVID-19 thromboinflammatory firestorm. JCI Insight 5, (2020).

68. M. G. Tektonidou, L. Andreoli, M. Limper, Z. Amoura, R. Cervera, N. Costedoat-Chalumeau, M. J. Cuadrado, T. Dorner, R. Ferrer-Oliveras, K. Hambly, M. A. Khamashta, J. King, F. Marchiori, P. L. Meroni, M. Mosca, V. Pengo, L. Raio, G. Ruiz-Irastorza, Y. Shoenfeld, L. Stojanovich, E. Svenungsson, D. Wahl, A. Tincani, M. M. Ward, EULAR recommendations for the management of antiphospholipid syndrome in adults. Ann Rheum Dis 78, 1296–1304 (2019).

69. D. G. Wahl, F. Guillemin, E. de Maistre, C. Perret-Guillaume, T. Lecompte, G. Thibaut, Meta-analysis of the risk of venous thrombosis in individuals with antiphospholipid antibodies without underlying autoimmune disease or previous thrombosis. Lupus 7, 15–22 (1998).

70. M. Galli, D. Luciani, G. Bertolini, T. Barbui, Lupus anticoagulants are stronger risk factors for thrombosis than anticardiolipin antibodies in the antiphospholipid syndrome: a systematic review of the literature. Blood 101, 1827–1832 (2003).

71. Q. Reynaud, J. C. Lega, P. Mismetti, C. Chapelle, D. Wahl, P. Cathebras, S. Laporte, Risk of venous and arterial thrombosis according to type of antiphospholipid antibodies in adults without systemic lupus erythematosus: a systematic review and meta-analysis. Autoimmun Rev 13, 595–608 (2014).

72. D. G. Wahl, F. Guillemin, E. de Maistre, C. Perret, T. Lecompte, G. Thibaut, Risk for venous thrombosis related to antiphospholipid antibodies in systemic lupus erythematosus- -a meta-analysis. Lupus 6, 467–473 (1997).

73. N. Heikal, T. B. Martins, S. K. White, R. Willis, D. Ware Branch, R. L. Schmidt, A. E. Tebo, Laboratory Evaluation of Antiphospholipid Syndrome. Am J Clin Pathol 152, 638–646 (2019).

74. G. R. Hughes, M. A. Khamashta, Seronegative antiphospholipid syndrome. Ann Rheum Dis 62, 1127 (2003).

75. N. J. Sweiss, R. Bo, R. Kapadia, D. Manst, F. Mahmood, T. Adhikari, S. Volkov, M. Badaracco, M. Smaron, A. Chang, J. Baron, J. S. Levine, IgA anti-beta2-glycoprotein I autoantibodies are associated with an increased risk of thromboembolic events in patients with systemic lupus erythematosus. PLoS One 5, e12280 (2010).

76. S. Sciascia, G. Sanna, V. Murru, D. Roccatello, M. A. Khamashta, M. L. Bertolaccini, Antiprothrombin (aPT) and anti-phosphatidylserine/prothrombin (aPS/PT) antibodies and the risk of thrombosis in the antiphospholipid syndrome. A systematic review. Thromb Haemost 111, 354–364 (2014).

77. C. A. Nunez-Alvarez, G. Hernandez-Molina, P. Bermudez-Bermejo, V. Zamora-Legoff, D. F. Hernandez-Ramirez, E. Olivares-Martinez, A. R. Cabral, Prevalence and associations of anti-phosphatidylserine/prothrombin antibodies with clinical phenotypes in patients with primary antiphospholipid syndrome: aPS/PT antibodies in primary antiphospholipid syndrome. Thromb Res 174, 141–147 (2019).

78. M. Ehrenfeld, A. Tincani, L. Andreoli, M. Cattalini, A. Greenbaum, D. Kanduc, J. Alijotas-Reig, V. Zinserling, N. Semenova, H. Amital, Y. Shoenfeld, Covid-19 and autoimmunity. Autoimmun Rev 19, 102597 (2020).

79. K. H. Lee, A. Kronbichler, D. D. Park, Y. Park, H. Moon, H. Kim, J. H. Choi, Y. Choi, S. Shim, I. S. Lyu, B. H. Yun, Y. Han, D. Lee, S. Y. Lee, B. H. Yoo, K. H. Lee, T. L. Kim, H. Kim, J. S. Shim, W. Nam, H. So, S. Choi, S. Lee, J. I. Shin, Neutrophil extracellular traps (NETs) in autoimmune diseases: A comprehensive review. Autoimmun Rev 16, 1160–1173 (2017).

80. I. Jeremic, O. Djuric, M. Nikolic, M. Vlajnic, A. Nikolic, D. Radojkovic, B. Bonaci-Nikolic, Neutrophil extracellular traps-associated markers are elevated in patients with systemic lupus erythematosus. Rheumatol Int 39, 1849–1857 (2019).

81. M. Steri, V. Orru, M. L. Idda, M. Pitzalis, M. Pala, I. Zara, C. Sidore, V. Faa, M. Floris, M. Deiana, I. Asunis, E. Porcu, A. Mulas, M. G. Piras, M. Lobina, S. Lai, M. Marongiu, V. Serra, M. Marongiu, G. Sole, F. Busonero, A. Maschio, R. Cusano, G. Cuccuru, F. Deidda, F. Poddie, G. Farina, M. Dei, F. Virdis, S. Olla, M. A. Satta, M. Pani, A. Delitala, E. Cocco, J. Frau, G. Coghe, L. Lorefice, G. Fenu, P. Ferrigno, M. Ban, N. Barizzone, M. Leone, F. R. Guerini, M. Piga, D. Firinu, I. Kockum, I. Lima Bomfim, T. Olsson, L. Alfredsson, A. Suarez, P. E. Carreira, M. J. Castillo-Palma, J. H. Marcus, M. Congia, A. Angius, M. Melis, A. Gonzalez, M. E. Alarcon Riquelme, B. M. da Silva, M. Marchini, M. G. Danieli, S. Del Giacco, A. Mathieu, A. Pani, S. B. Montgomery, G. Rosati, J. Hillert, S. Sawcer, S. D’Alfonso, J. A. Todd, J. Novembre, G. R. Abecasis, M. B. Whalen, M. G. Marrosu, A. Meloni, S. Sanna, M. Gorospe, D. Schlessinger, E. Fiorillo, M. Zoledziewska, F. Cucca, Overexpression of the Cytokine BAFF and Autoimmunity Risk. N Engl J Med 376, 1615-1626 (2017).

82. C. Lood, G. C. Hughes, Neutrophil extracellular traps as a potential source of autoantigen in cocaine-associated autoimmunity. Rheumatology (Oxford) 56, 638–643 (2017).

83. S. W. Jackson, A. Davidson, BAFF inhibition in SLE-Is tolerance restored? Immunol Rev 292, 102–119 (2019).

84. S. Bucciarelli, G. Espinosa, R. Cervera, D. Erkan, J. A. Gomez-Puerta, M. Ramos-Casals, J. Font, R. A. Asherson, A. European Forum on Antiphospholipid, Mortality in the catastrophic antiphospholipid syndrome: causes of death and prognostic factors in a series of 250 patients. Arthritis Rheum 54, 2568–2576 (2006).

85. Y. Zuo, S. Yalavarthi, H. Shi, K. Gockman, M. Zuo, J. A. Madison, C. N. Blair, A. Weber, B. J. Barnes, M. Egeblad, R. J. Woods, Y. Kanthi, J. S. Knight, Neutrophil extracellular traps in COVID-19. JCI Insight, (2020).

86. D. Erkan, M. D. Lockshin, A. A. members, APS ACTION--AntiPhospholipid Syndrome Alliance For Clinical Trials and InternatiOnal Networking. Lupus 21, 695–698 (2012).

87. S. Sciascia, R. Willis, V. Pengo, S. Krilis, D. Andrade, M. G. Tektonidou, A. Ugarte, C. Chighizola, D. W. Branch, R. A. Levy, C. Nalli, P. R. Fortin, M. Petri, E. Rodriguez, I. Rodriguez-Pinto, T. Atsumi, I. Nascimento, R. Rosa, A. Banzato, D. Erkan, H. Cohen, M. Efthymiou, I. Mackie, M. L. Bertolaccini, A. Aps, The comparison of real world and core laboratory antiphospholipid antibody ELISA results from antiphospholipid syndrome alliance for clinical trials & international networking (APS ACTION) clinical database and repository analysis. Thromb Res 175, 32–36 (2019).

88. G. Lakos, E. J. Favaloro, E. N. Harris, P. L. Meroni, A. Tincani, R. C. Wong, S. S. Pierangeli, International consensus guidelines on anticardiolipin and anti-beta2-glycoprotein I testing: report from the 13th International Congress on Antiphospholipid Antibodies. Arthritis Rheum 64, 1–10 (2012).

89. K. Kessenbrock, M. Krumbholz, U. Schonermarck, W. Back, W. L. Gross, Z. Werb, H. J. Grone, V. Brinkmann, D. E. Jenne, Netting neutrophils in autoimmune small-vessel vasculitis. Nat Med 15, 623–625 (2009).

90. V. Yadav, L. Chi, R. Zhao, B. E. Tourdot, S. Yalavarthi, B. N. Jacobs, A. Banka, H. Liao, S. Koonse, A. C. Anyanwu, S. H. Visovatti, M. A. Holinstat, J. M. Kahlenberg, J. S. Knight, D. J. Pinsky, Y. Kanthi, Ectonucleotidase tri(di)phosphohydrolase-1 (ENTPD-1) disrupts inflammasome/interleukin 1beta-driven venous thrombosis. J Clin Invest 129, 2872–2877 (2019).

91. O. R. Palmer, M. E. Shaydakov, J. P. Rainey, D. A. Lawrence, J. M. Greve, J. A. Diaz, Update on the electrolytic IVC model for pre-clinical studies of venous thrombosis. Res Pract Thromb Haemost 2, 266–273 (2018).

92. D. A. Slatter, C. L. Percy, K. Allen-Redpath, J. M. Gajsiewicz, N. J. Brooks, A. Clayton, V. J. Tyrrell, M. Rosas, S. N. Lauder, A. Watson, M. Dul, Y. Garcia-Diaz, M. Aldrovandi, M. Heurich, J. Hall, J. H. Morrissey, S. Lacroix-Desmazes, S. Delignat, P. V. Jenkins, P. W. Collins, V. B. O’Donnell, Enzymatically oxidized phospholipids restore thrombin generation in coagulation factor deficiencies. JCI Insight 3, (2018).

